# A genome-wide association study of COVID-19 related hospitalization in Spain reveals genetic disparities among sexes

**DOI:** 10.1101/2021.11.24.21266741

**Authors:** Ángel Carracedo, Spanish COalition to Unlock Research on host GEnetics on COVID-19 (SCOURGE)

**Affiliations:** University of Santiago de Compostela

## Abstract

We describe the results of the Spanish Coalition to Unlock Research on Host Genetics on COVID-19 (SCOURGE). In sex-disaggregated genome-wide studies of COVID-19 hospitalization, we found two known loci associated among males (*SLC6A20*-*LZTFL1* and *IFNAR2*), and a novel one among females (*TLE1*). Meta-analyses with independent studies revealed two novel associations (*AQP3* and *ARHGAP33*) and replicated *ELF5*. A genetic risk score predicted COVID-19 severity, especially among younger males. We found less SNP-heritability and larger heritability differences by age (<60/≥60 years) among males than females. Inbreeding depression was associated with COVID-19 hospitalization and severity, and the effect was stronger among older males.

## Introduction

Coronavirus disease 2019 (COVID-19), caused by the severe acute respiratory syndrome coronavirus 2 (SARS-CoV-2), develops with wide clinical variability, ranging from asymptomatic infection to a life-threatening condition [1]. Advanced age and the presence of comorbidities are well-known major risk factors of COVID-19 severity [2, 3]. In addition, male sex is another risk factor associated with COVID-19 severity, regardless of comorbidities [4].

International genetic studies based on genome-wide association studies (GWAS) and/or comparative genome sequencing analyses have been conducted to identify genetic variants associated with COVID-19 severity [5, 6]. These studies have revealed the role of genes of the Type-I interferon (IFN) signalling pathway as key players underlying disease severity [7–9]. Besides, they have also identified other potential loci previously linked to lung function and diseases and autoimmunity [9]. Regarding COVID-19 severity in males, sex-disaggregated genetic analyses have received limited attention despite the importance of sex disparities in clinical severity [10]. Early studies suggested immune deficits presumably due to pre-existing neutralizing autoantibodies against Type-I IFN in older male patients [11].

The effects of autozygosity, measured as the change of the mean value of a complex trait due to inbreeding, have been useful to identify alternative genetic risk explanations and effects that traditionally are not captured by GWAS [12]. By analysing the contribution of the inbreeding depression (ID) through the lens of the runs of homozygosity (ROH: genomic tracts where homozygous markers occur in an uninterrupted sequence), it is possible to assess the importance of directional dominance or overdominance in the genetic architecture of complex traits [13]. Even though this is a relatively modern approach, different studies have shown the importance of homozygosity in a large range of complex phenotypes, including anthropometric, cardiometabolic, and mental traits [14–16].

Through diverse nested sub-studies, the Spanish Coalition to Unlock Research on Host Genetics on COVID-19 (SCOURGE) consortium was launched in May 2020 aiming to find biomarkers of evolution and prognosis that can have an immediate impact on clinical management and therapeutic decisions in SARS-CoV-2 infections. This consortium has recruited patients from hospitals across Spain and Latin America in close collaboration with the STOP-Coronavirus initiative (https://www.scourge-covid.org). Here we describe the results of the first SCOURGE genome-wide studies of COVID-19 conducted in patients recruited in Spain. To the best of our knowledge, this is the first time that the impact of homozygosity is considered in COVID-19 studies, serving as a complement to the traditional GWAS to assess both the additive and dominant components of the genetic architecture of COVID-19 severity. Likewise, the ID analysis could also help to explain the strong effect of age in COVID-19 severity.

## Results

### Discovery phase

In the SCOURGE study, 11,939 COVID-19 positive cases were recruited from 34 centres (**Supplementary Table 1**). All diagnosed cases were classified in a five-level severity scale (**Extended Data Table 1**). Two untested Spanish sample collections were used as general population controls in some analyses: 3,437 samples from the Spanish DNA biobank (https://www.bancoadn.org) and 2,506 samples from the GR@CE consortium [17]. The discovery phase samples were genotyped with the Axiom Spain Biobank Array (Thermo Fisher Scientific). Details of quality control, ancestry inference and imputation are shown in the Methods section. Individuals with inferred European ancestry were used for association testing. After post-imputation filtering, 15,045 individuals (9,371 COVID-19 positive cases and 5,674 population controls) and 8,933,154 genetic markers remained in the SCOURGE European study (discovery). Clinical and demographic characteristics of European patients from SCOURGE included in the analysis are shown in **Extended Data Table 2**.

The discovery phase of the GWAS was carried out with infection susceptibility and three severity outcomes (hospitalization, severe illness, and critical illness) which were tested using three different control definitions (see **Supplementary Table 2**).

- A1 analysis: COVID-19 positive not satisfying the case condition and control samples from the general population (COVID-19 untested).

- A2 analysis: control samples from the general population.

- C analysis: COVID-19 positive not satisfying the case condition.

The GWAS was carried on by fitting logistic mixed regression models adjusting for age, sex, and the first 10 Principal Components (PCs) (**Methods**). **Supplementary Table 3** shows the independent significant associated loci for hospitalization, severity, critical illness, and infection susceptibility risk, for global and sex-stratified analysis in the SCOURGE dataset. However, considering the overlap between the findings for these analyses, only the main results for the A1 analysis are presented. The SCOURGE Board of Directors has agreed to aggregate the GWAS summaries with those from the COVID-19 Host Genetics Initiative (HGI).

All analyses support the association of two known loci, 3p21.31 and 21q22.11. However, other suggestive associations were also found (**Figure 1** and **Extended Data Figure 1**). Strikingly, the leading signals found in the global (sex-aggregated) analysis were genome-wide significant in the analyses among males but not among females. However, the leading variant of 9q21.32 (near *TLE1* gene) reached genome-wide significance among females only. Several variants (rs17763742 near *LZTFL1*, rs2834164 in *IFNAR2*, and rs1826292621 near *TLE1*) showed a significant difference in effect sizes (SNP*sex interaction *p*<0.0031, adjusted probability for 16 comparisons) linked not only to hospitalization, but also to critical illness and infection risk. The A2 and C analyses did not reveal any additional significant loci (**Supplementary Figure 1**). While fine-mapping studies in 3p21.31 and 21q22.11 have led to gene and variant prioritization within these loci (**Extended Data Figure 2**), a Bayesian fine-mapping on the 9q21.32 did not allow to prioritize variants by their role as expression quantitative trait loci (eQTLs) or anticipate the function (**Figure 2**). To assess if a higher prevalence of comorbidities in males may underlie these differential findings, the presence of comorbidities was tested for association within hospitalized patients. No significant association was found in either males or females (**Supplementary Figure 2**). Further explorations of the genetic associations with comorbidities are presented in the **Supplementary Note**.

**Figure 1.**
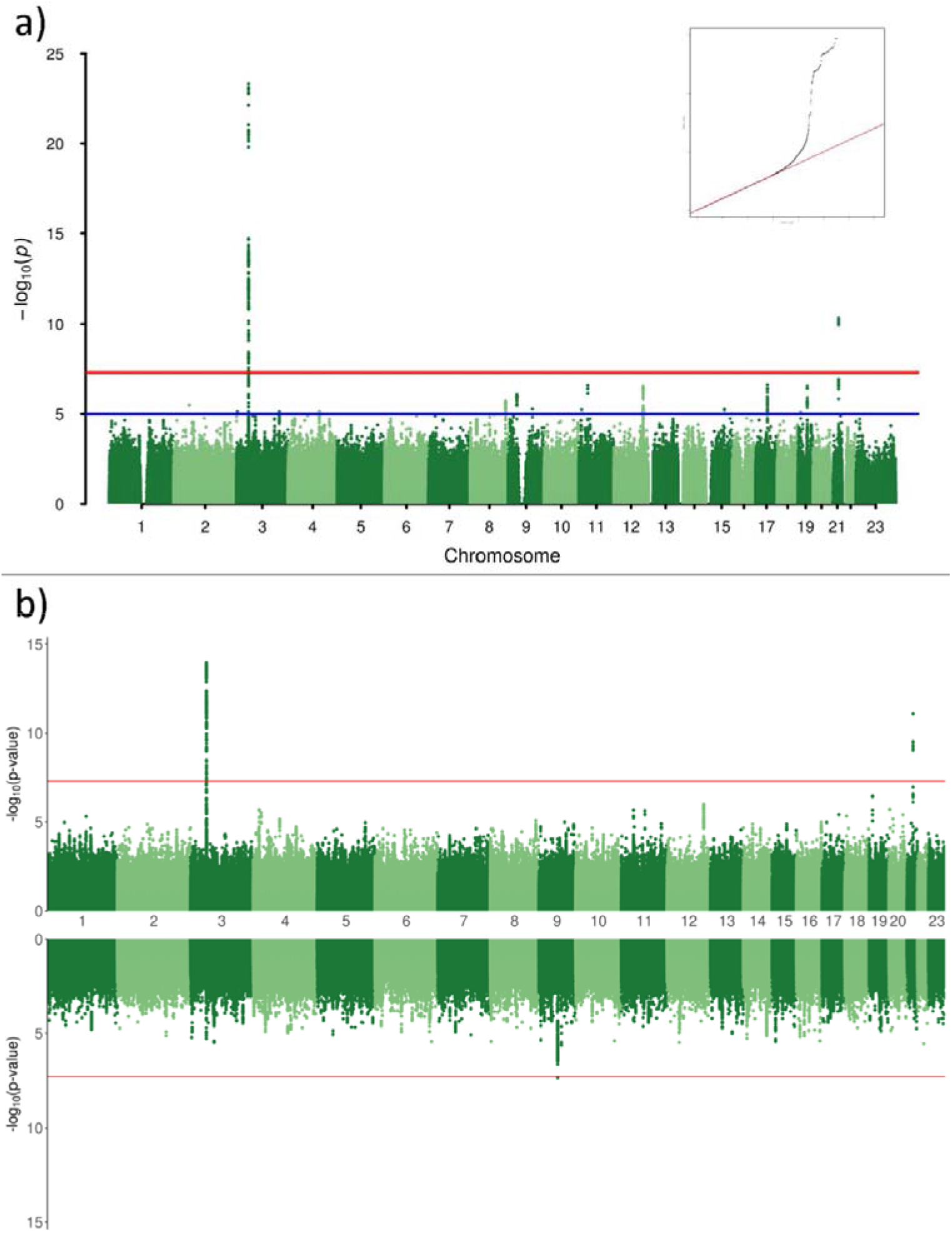
Association results of SCOURGE for A1 hospitalization analysis in a) global analysis, and b) sex-disaggregated analyses (Miami plot, top panel: males, bottom panel: females). A quantile-quantile plot of the global analysis is also shown as an inset.

**Figure 2.**
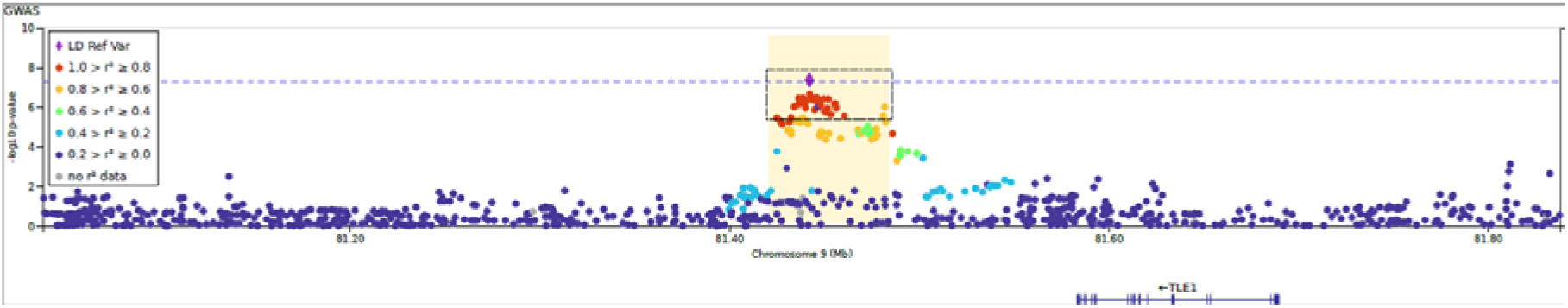
Regional plot of a novel association at 9q21.32 found among females from the SCOURGE study. The x axis reflect the chromosomal position, and the y axis the -log(*p*-value). The sentinel variant is indicated by a diamond and all other variants are colour coded by their degree of linkage disequilibrium with the sentinel variant in Europeans. Credible set for this signal is shown within a dashed square. The horizontal dotted blue line corresponds to the threshold for genome-wide significance (*p*=5×10^-8^).

This GWAS phase was also performed disaggregated by age (<60/≥60 years old), and by age and sex simultaneously. Differences in effect sizes between both age groups were tested for the SNPs shown in the **Supplementary Table 3**, in global and sex-specific analysis (**Supplementary Table 4**). Significant findings were only found in the subgroup of males with <60 years old. Differences in effect size (significant age-interaction) were significant at 3p21.31 for severity and critical illness, and suggestive in hospitalization.

### Lookup of previously found COVID-19 host risk factors in the SCOURGE study

Known significant loci for COVID-19 severity in 3p21.31 (near *SLC6A20* and *LZTFL1*) and 21q22.11 (in *IFNAR2*) were clearly replicated at genome-wide significance in this study, specifically for risk of infection, hospitalization, and severity risk. Three other loci, in 9q34.2 (in *ABO*), 12q24.13 (in *OAS1*), and 19p13.2 (near *RAVER1* and *TYK2*), did not reach the genome-wide significance threshold but they were significant after correcting for the 390 tests performed in a lookup (13 SNPs and 30 analyses, significance threshold *p*<1.3×10^-4^). In agreement with previous results, *ABO* was mainly associated with the risk of infection. However, other loci as those in 3q12.3 (near *RPL24*) and 19p13.3 (near *DPP9*), previously found associated with COVID-19 severity, were not replicated in the SCOURGE Europeans. The complete list of results for the list of COVID-19 HGI significant loci [9] is shown in **Figure 3a** and in the **Supplementary Table 5**. **Figure 3a** also reinforces the clear sex differences found in this study.

**Figure 3.**
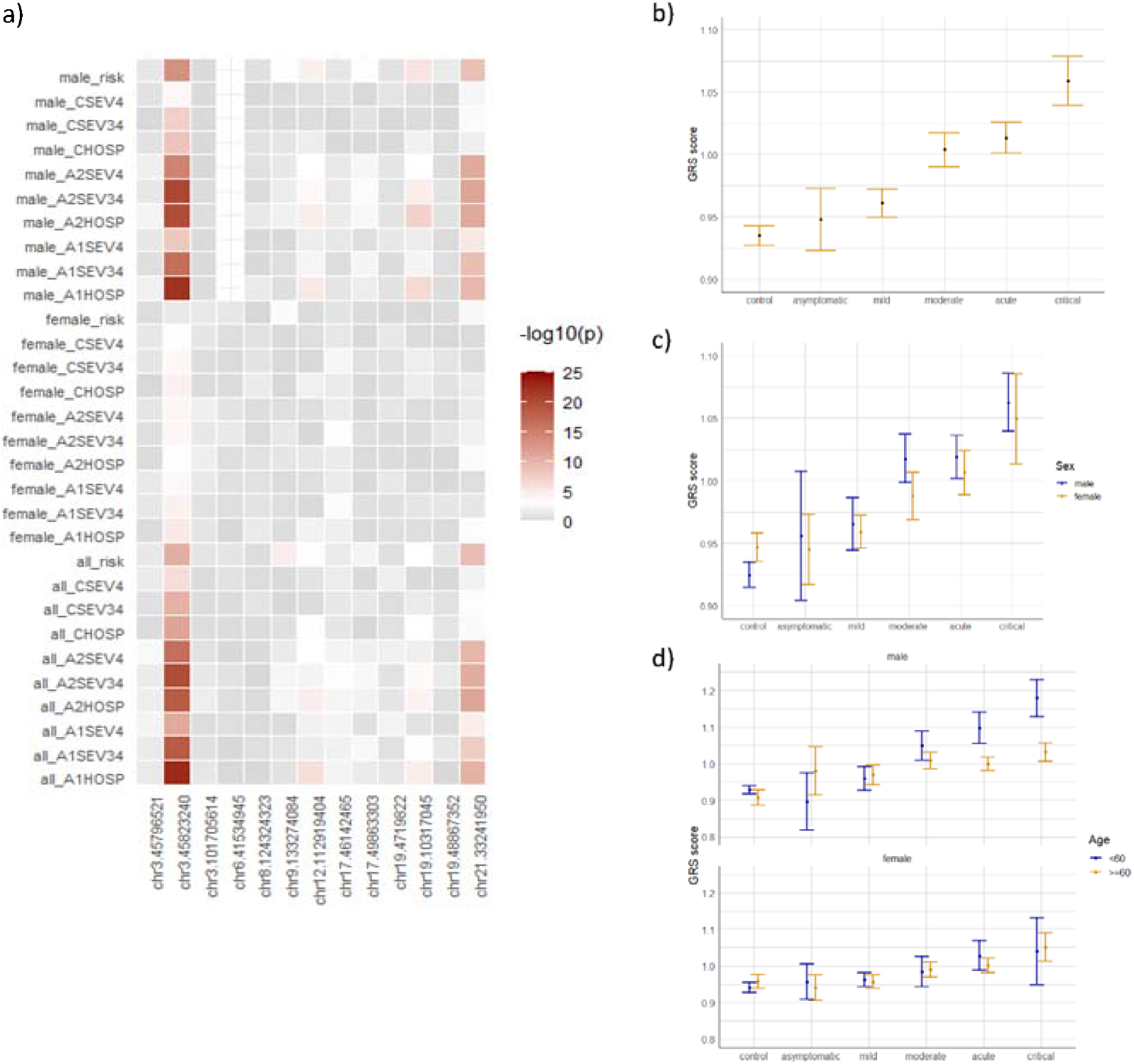
a) Heatmap illustrating the results in all analyses performed in this study (rows) for the 13 leading variants in the COVID-19 HGI study (columns). Each box illustrates the top associated variant within the focal region. The colour (grey to dark red) indicates the strength (significance level) of the association in SCOURGE. Note: In three cases (chr12: 112919388, chr19: 4719431 and chr21: 33242905), the imputed variants did not pass QC filters in SCOURGE and they were replaced by the nearest QC-ed imputed variant (chr12:112919404, chr19:4719822, and chr21:33241950, respectively). b-d) Estimates of the GRS mean (and 95% confidence interval) built from the 13 leading variants found by the COVID-19 HGI GWAS for each category of the severity scale recorded in SCOURGE in global (b), sex-disaggregated (c) and sex-age disaggregated analysis (d).

### Genotype risk score and the COVID-19 severity scale

We developed a GRS combining the 13 leading variants found by the COVID-19 HGI GWAS to appraise its prediction power of the severity scale in SCOURGE. The average values of the GRS for each of the severity scale levels of SCOURGE were statistically different between the six levels in global (*F*_5,14547_=50.7, *p*<2×10^-16^) and the sex-stratified analyses (females: *F*_4,4753_=10.30, *p=*2.62×10^-8^; males: *F*_4,4114_=10.47, *p=*1.94×10^-8^) (**Figure 3b, 3c**). Duncan’s *post hoc* test did not support differentiation between some of the severity levels, roughly defining three classes: one comprising controls and the asymptomatic and mild cases; another with moderate and severe cases; and one with the critical cases. Within each category, we did not find any statistically significant differences between sexes, yet interestingly the GRS mean remained higher for males than for females while this trend was reversed in the control group (**Figure 3b**). Moreover, the GRS mean was not equal for both sexes (*t*_8994.5_=-5.21, *p=*1.98×10^-7^). When the GRS was performed disaggregating by age (<60/≥60 years old) and sex simultaneously, we found the same three severity classes in the subgroup of males with <60 years old, supporting the importance of this group in the overall findings (**Figure 3c**). In fact, significant differences were found between both age groups within males with severe (*t*_359.2_ = 4.18, *p*= 3.6×10^-5^) and critical illness (*t*_815_ = 5.12, *p*= 3.9×10^-7^).

### Replication phase

Results for hospitalization risk were meta-analysed with a second Spanish cohort, the CNIO study (**Methods**). This study was filtered following the same quality control and imputation procedures. The final dataset of the CNIO study included 2,446 European individuals (1,378 COVID-19 positive cases and 1,068 population controls) and 8,895,721 markers.

**Table 1** shows the results that were genome-wide significant either in global or sex-stratified meta-analysis with SCOURGE. Besides the widely replicated loci at 3p21.31 and 21q22.11, three additional signals were found: chr9:33426577:A:G (intergenic to *AQP7* and *AQP3*), chr17:45422978:G:C (intronic to *ARHGAP27*), and chr19:35687796:G:A (intergenic to *UPK1A* and *ZBTB32*). Bayesian fine-mapping around chr17:45422978:G:C failed to prioritize a credible set of variants, hindering functional links of the locus. Functional assessments of the prioritized variants by the Bayesian fine-mapping analysis in the other two regions supported that they were eQTLs of the *AQP3* and *ARGAP33* genes, including in whole blood and lung tissues (**Figure 4**).

**Table 1.** Genome-wide significant variants in global or sex-stratified meta-analysis between the SCOURGE and CNIO studies. Representative SNPs were selected using the clump function of PLINK 1.9 (clumping parameters r^2^=0.5, pval=5×10^-8^ and pval_2_=0.05).

| SNP | chr:position | EA | NEA | Meta-ALL |  |  | Meta-males |  |  | Meta-females |  |  | Nearest gene |
| --- | --- | --- | --- | --- | --- | --- | --- | --- | --- | --- | --- | --- | --- |
|  |  |  |  | beta | SE | p-value | beta | SE | p-value | beta | SE | p-value |  |
| rs115679256 | 3:45587795 | G | A | 0.43 | 0.08 | 1.1E-08 | 0.48 | 0.10 | 2.3E-06 | 0.40 | 0.11 | 2.9E-04 | LIMD1 |
| rs17763742 | 3:45805277 | A | G | 0.60 | 0.05 | 4.1E-29 | 0.74 | 0.07 | 3.3E-25 | 0.43 | 0.08 | 4.5E-08 | LZTFL1 |
| rs35477280 | 3:45932600 | G | A | 0.39 | 0.05 | 1.4E-17 | 0.48 | 0.06 | 6.3E-15 | 0.28 | 0.07 | 1.6E-05 | FYCO1 |
| rs4443214 | 3:46136372 | T | C | 0.25 | 0.04 | 9.0E-09 | 0.26 | 0.06 | 1.4E-05 | 0.26 | 0.06 | 4.4E-05 | XCR1 |
| rs115102354 | 3:46180545 | A | G | 0.41 | 0.07 | 1.6E-08 | 0.52 | 0.10 | 2.1E-07 | 0.32 | 0.10 | 2.0E-03 | CCR3 |
| rs10813976 | 9:33426577 | A | G | 0.18 | 0.03 | 2.7E-08 | 0.16 | 0.04 | 2.5E-04 | 0.19 | 0.05 | 3.5E-05 | AQP3 |
| rs1230082 | 17:45422978 | C | G | 0.16 | 0.03 | 2.1E-08 | 0.17 | 0.04 | 2.8E-05 | -0.15 | 0.04 | 2.5E-04 | ARHGAP27 |
| rs77127536 | 19:35687796 | G | A | -0.22 | 0.04 | 1.3E-08 | -0.25 | 0.05 | 2.1E-06 | -0.19 | 0.05 | 4.3E-04 | UPK1A/ZBTB32 |
| rs17860169 | 21:33240996 | A | G | 0.19 | 0.03 | 2.3E-11 | 0.27 | 0.04 | 1.4E-11 | 0.12 | 0.04 | 3.7E-03 | IFNAR2 |

EA=Effect Allele; NEA=Non-Effect Allele; beta=Effect coefficient; SE=Standard Error

**Figure 4.**
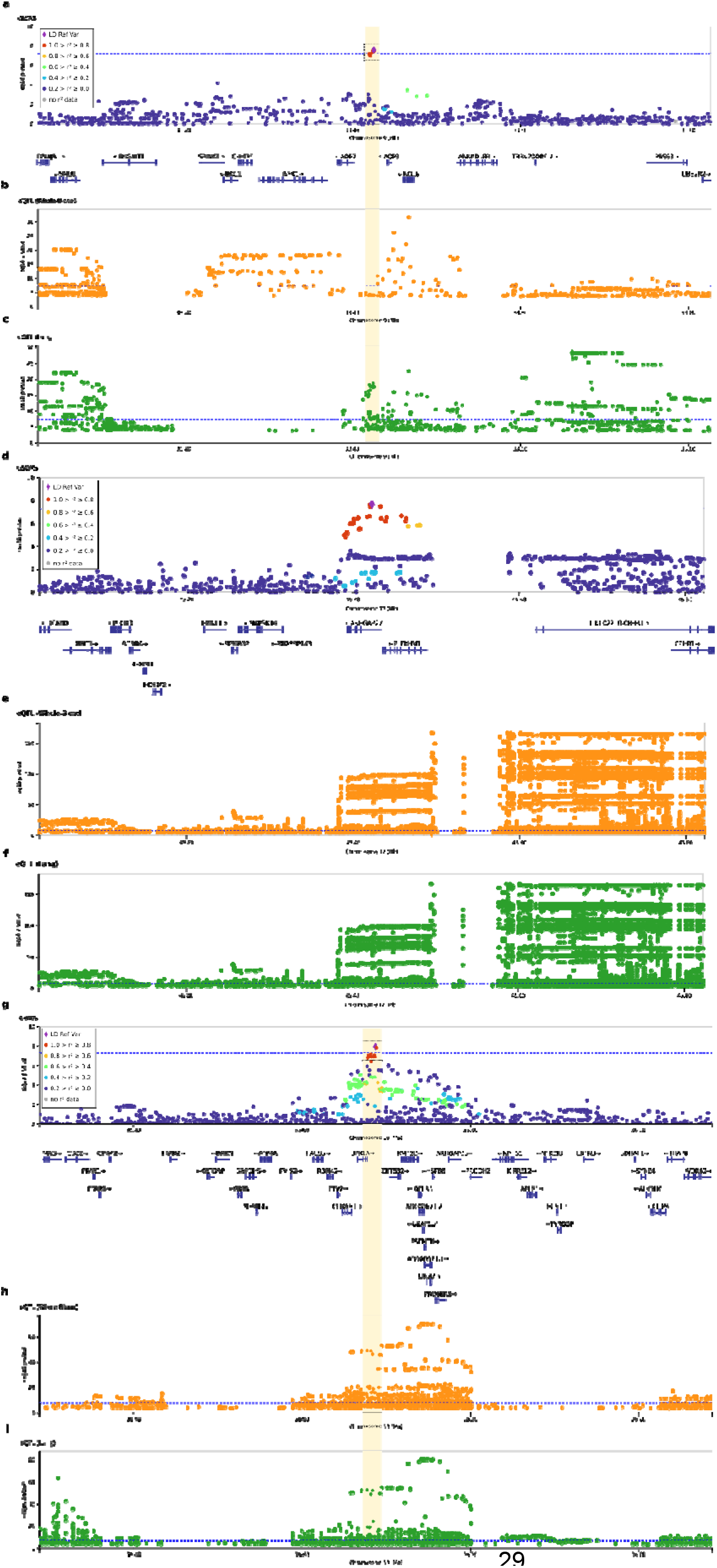
Regional plots of novel association signals found in 9p13.3 (a-c), 17q21.31 (d-f), and 19q13.12 (g-i) from the meta-analysis between the SCOURGE and CNIO studies. The x axes reflect the chromosomal position, and the y axes the -log(*p*-value) of the SCOURGE-CNIO meta-analysis. On panels a, d, and g the sentinel variant is indicated by a diamond and all other variants are colour coded by their degree of linkage disequilibrium with the sentinel variant in Europeans. Whenever a concise set of variants was prioritized, a credible set is shown within a dashed square. The horizontal dotted blue line corresponds to the threshold for genome-wide significance (*p*=5×10^-8^). In the rest of panels, the x axes reflect the chromosomal position, and the y axes the -log(*p*-value) resulting from the eQTL analyses in whole blood (b, e, and h) and in the lung (c, f, and i) whenever a significant finding is available from GTEx v8.

These variants were also associated with the three severity groups previously outlined in SCOURGE by the GRS under a multinomial model (**Supplementary Table 6**).

### Validation of results in independent European studies

Hospitalization risk was meta-analysed with other European studies combining both Spanish cohorts (SCOURGE and CNIO) with four other sex-disaggregated studies from the COVID-19 HGI consortium, namely: BelCOVID, GenCOVID, Hostage-Spain, and Hostage-Italy (**Table 2**). Once again, the most outstanding significant loci were found at 3p21.31 and 21q22.11 (in global and male-specific analyses), and three additional loci reached genome-wide significance in the meta-analysis of males: chr12:11292383:A:G (in *OAS1*), chr19:35687796:G:A (intergenic to *UPK1A* and *ZBTB32*), and chr11:34482745:G:A (in *ELF5*). The 3p21.31 variants reached genome-wide significance in females, although at significantly lower level than in males despite the similar sample sizes (Z=3.33, *p=*5×10^-4^).

**Table 2.** Results of European meta-analysis for hospitalization risk. Summary statistics of both phases (SCOURGE and CNIO) were meta-analysed with four additional sex-disaggregated European studies from the COVID-19 HGI consortium.

| SNP | chr:position | EA | NEA | Meta-all |  |  | Meta-males |  |  | Meta-females |  |  | Nearest gene |
| --- | --- | --- | --- | --- | --- | --- | --- | --- | --- | --- | --- | --- | --- |
|  |  |  |  | beta | SE | p-value | beta | SE | p-value | beta | SE | p-value |  |
| rs115679256 | 3:45587795 | G | A | 0.37 | 0.06 | 1.3E-08 | 0.41 | 0.08 | 5.6E-07 | 0.36 | 0.09 | 1.6E-04 | LIMD1 |
| rs13078854 | 3:45820440 | G | A | 0.53 | 0.04 | 6.7E-34 | 0.64 | 0.05 | 2.7E-33 | 0.38 | 0.06 | 1.0E-09 | LZTFL1 |
| rs41289622 | 3:45973053 | T | G | 0.36 | 0.04 | 3.6E-21 | 0.44 | 0.05 | 3.4E-20 | 0.27 | 0.05 | 7.2E-07 | FYCO1 |
| rs115102354 | 3:46180545 | A | G | 0.40 | 0.06 | 8.9E-12 | 0.48 | 0.07 | 6.8E-11 | 0.26 | 0.08 | 1.8E-03 | XCR1 |
| rs61882275 | 11:34482745 | G | A | -0.12 | 0.02 | 1.0E-06 | -0.17 | 0.03 | 4.1E-08 | -0.08 | 0.03 | 1.3E-02 | ELF5 |
| rs4767028 | 12:112921383 | A | G | -0.16 | 0.02 | 1.6E-10 | -0.19 | 0.03 | 2.5E-09 | -0.11 | 0.04 | 8.7E-04 | OAS1 |
| rs12609134 | 19:35687796 | G | A | -0.19 | 0.03 | 2.3E-08 | -0.22 | 0.04 | 9.5E-08 | -0.13 | 0.05 | 6.0E-03 | UPK1A/ZBTB32 |
| rs17860169 | 21:33240996 | A | G | 0.18 | 0.03 | 3.9E-12 | 0.21 | 0.03 | 1.6E-10 | 0.15 | 0.04 | 2.9E-05 | IFNAR2 |

EA=Effect Allele; NEA=Non-Effect Allele; beta=Effect coefficient; SE=Standard Error

Significance of two interesting loci revealed in the Spanish studies was reduced in the meta-analysis with other European studies, although still showing suggestive associations: that of 9q21.32 near *TLE1* previously found only in females (female meta-analysis beta=0.29 *p=*5.4×10^-7^), and that of 9p13.3 near *AQP3* (global meta-analysis beta=0.15, *p=*1.23×10^-7^).

### Heritability of COVID-19 hospitalization

In the hospitalization risk analysis, we found that common variants (MAF >1%) explain 27.1% (95%CI: 19.0-35.3%) of heritability on the observed scale (corresponding to 13.1% [95%CI: 9.2-17.0%] on the liability scale, assuming a prevalence of 0.5%) (**Figure 5**). We observed less heritability among males than females (2.9% [95%CI: 0.00-10.6%] in males and 17.0% [95%CI: 9.2-24.9%] in females on the liability scale), which is in agreement with their higher risk of severe COVID-19 and with the observations that non-genetic factors (e.g. IFN autoantibodies) causing critical COVID-19 are more prevalent among males than females [11, 18]. In agreement with this idea, we observed larger heritability differences by age groups among males (40.2% [95%CI: 22.8-57.5%] in <60 years vs. 17.6% [95%CI: 0.00-38.0%] in ≥60 years on the liability scale) than among females (9.1% [0.00-31.3%] in <60 years vs. 13.7% [0.00-29.6%] in ≥60 years on the liability scale). This observation might be explained by the presence of X-linked deleterious variants such as those described in the *TLR7* gene that are life-threatening for COVID-19 among males [19–21].

**Figure 5.**
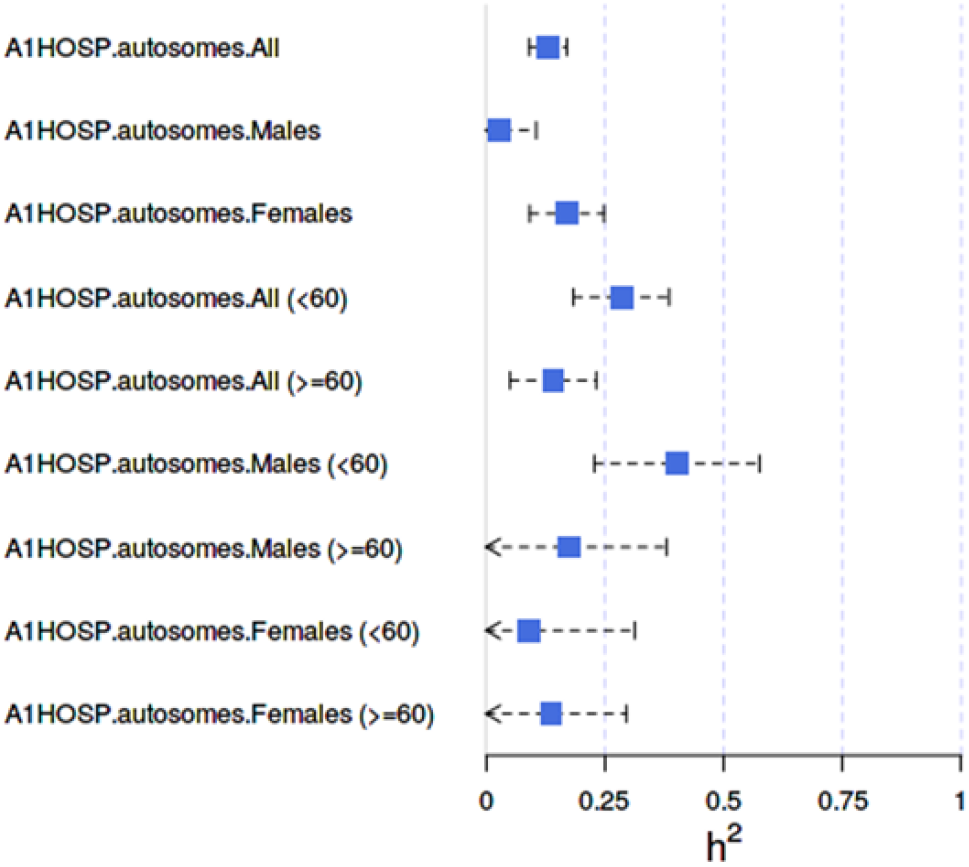
Forest plot of the SNP heritability estimates for the COVID-19 hospitalization risk analysis on the liability scale.

### Inbreeding depression and COVID-19 outcomes

ROH calling was performed in the European QC-ed genotyped dataset. Inbreeding depression (ID) analyses are described in **Methods** section and **Supplementary Note**.

The median genomic inbreeding coefficient, F_ROH_, for the entire SCOURGE study was 0.0048 (IQR = 0.004). No differences were detected between males (F_ROH_ = 0.004, IQR = 0.0035) and females (F_ROH_ = 0.0056, IQR = 0.0038), or between younger and older individuals (F_ROH individuals < 60 years old_ = 0.004, IQR = 0.0035; F_ROH individuals ≥ 60 years old_ = 0.0052, IQR = 0.0047, respectively) (**Supplementary Figure 3**). Regarding the ID in COVID-19 outcomes, we detected a positive association of the F_ROH_ in COVID-19 hospitalization risk (**Figure 6**), severity risk, and risk for critical illness (**Supplementary Table 7**). Our results showed that the hospitalization odds for COVID-19 patients with an F_ROH_ = 0.0039 were 380% higher than individuals with F_ROH_ = 0. No effect of the genomic relationship matrix (F_GRM_) was found.

**Figure 6.**
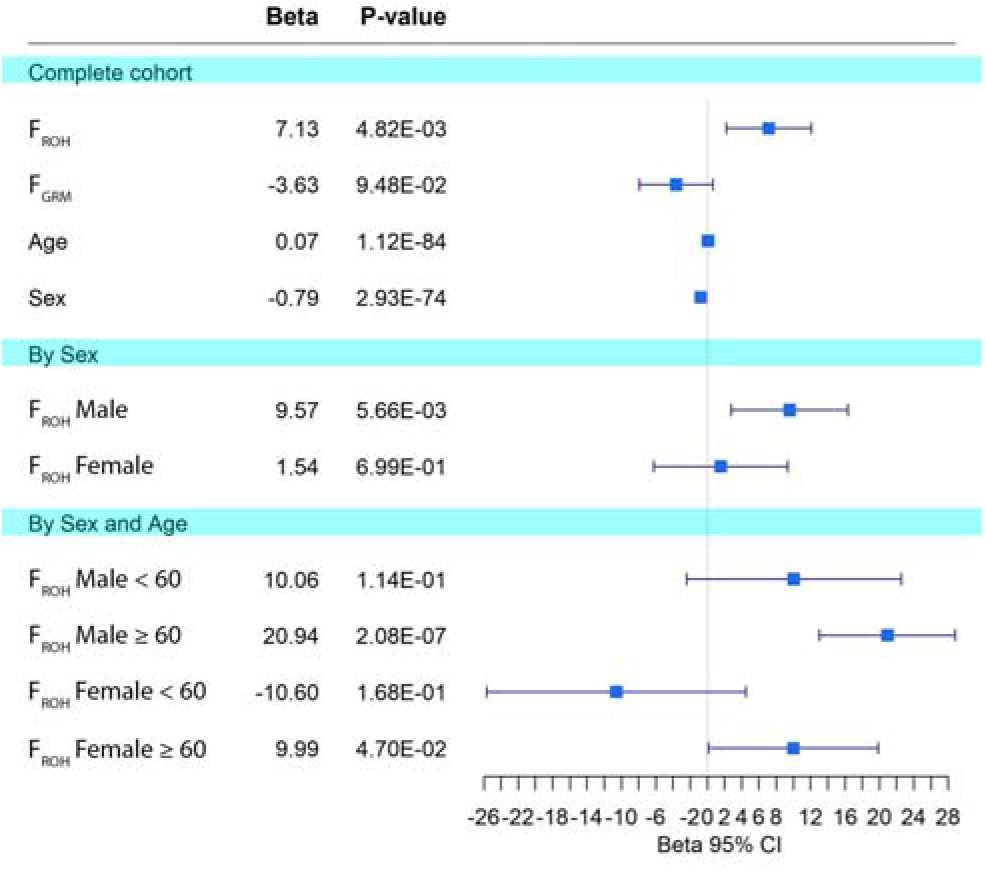
Effect of the inbreeding depression on COVID-19 hospitalization in the SCOURGE cohort. Forest plots are shown for global analyses as well as for sex and age-disaggregated analyses.

To assess whether ID in COVID-19 hospitalization in SCOURGE was different between sexes, we tested first the interaction between F_ROH_ and biological sex. F_ROH_, sex and the interaction of both (F_ROH_:Sex) were significant (F_ROH_ = 4.7×10^-3^, sex = 1.0×10^-112^, F_ROH_:Sex = 1.2×10^-3^). This interaction was significant when comparing the hospitalized COVID-19 patients with different controls (A2 and C analyses, see **Supplementary Table 8**). This interaction was also found with severity risk, but not with risk for critical illness (**Supplementary Table 8**). In sex-disaggregated analyses, we observed a sex-specific effect of inbreeding. F_ROH_ was significant in hospitalized males but not in females (**Figure 6** and **Supplementary Table 8**). This sex-specific effect was also observed with severity risk and in risk for critical illness (**Supplementary Table 8**). We then assessed whether ID in COVID-19 hospitalization was different by age. We detected a significant interaction between age and F_ROH_ for the three outcomes considered (hospitalization risk, severity risk, and critical illness risk) (**Supplementary Table 9**). Disaggregating SCOURGE by sex and age (<60, ≥60) we found that the ID for hospitalization and severity risk were detected mainly in older males (**Figure 6** and **Supplementary Table 9**). We detected ID for hospitalization and severity in males, older than 60 years old, but it was marginally significant in females (**Figure 6** and **Supplementary Table 9**). Age and sex-specific effects in hospitalization risk and severity risk were robust across different experimental designs using different control groups (**Supplementary Figure 4**).

Finally, we then aimed to replicate the ID results with hospitalization risk, assessing sex and age-specific effects, in a 4,418 case-enriched European cohort made of 16 studies from nine countries. Median F_ROH_ in this other European cohort was slightly higher than that of SCOURGE: 0.05 (0.009 – 0.0577). A positive and significant ID in COVID-19 hospitalization was detected in this other European cohort when the entire cohort was considered (F_ROH_ Beta = 18.22, *p* = 3.33×10^-3^). F_GRM_ was not significant (F_GRM_ Beta = -7.34, *p* = 0.240). ID was also detected in hospitalized COVID-19 males but not in females (Male F_ROH_ Beta = 16.22, *p* = 3.31×10^-3^; Female F_ROH_ Beta = 15.65, *p* =0.269). F_GRM_ was not significant in males or in female analyses. When disaggregating by age, it was possible to detect ID in hospitalization only in males ≥60 years old (F_ROH_ Beta = 36.16, *p* = 3.34×10^-3^) (**Supplementary Table 10**).

No evidence was found of major loci that may be exerting large effects. Rather, polygenicity seems to underlie the ID association. Different ROHi and regions of heterozygosity (RHZ) were found to be unique for hospitalized COVID-19 individuals (males and females) and non-hospitalized males respectively (**Supplementary Note, Supplementary Table 11**). An enrichment analysis of pathways based on the protein coding genes present in ROH islands were also different between sexes (**Supplementary Note, Supplementary Table 12**), revealing links with coagulation and complement pathways in males.

## Discussion

Here we report the replication of six COVID-19 loci across analyses and provide evidence supporting four additional loci, two of them specifically detected in one sex (one of them among females and the other among males). Besides, our analyses provide new insights into disease severity disparities by sex and age and support the necessity of similarly stratified studies to increase the possibility of detecting additional risk variants. Our GWAS constitutes the largest study on COVID-19 genetic risk factors conducted in Spain, with an intrinsic design benefit that SCOURGE includes detailed clinical and genetic information gathered homogeneously across the country. Besides, the study included patients from the whole spectrum of COVID-19 severity covering from asymptomatic to life-threatening disease. To date, most research on COVID-19 disease has focused on respiratory failure. However, the inclusion of a severity scale provided a unique opportunity to assess whether previously reported loci combined into a GRS model were associated with differential risk by strata. Association was tested for four main variables: infection, hospitalization, severe illness, and critical illness, and using different definitions of controls to align with the COVID-19 HGI. Irrespective of the tested outcomes or the definition of controls, the results were very similar, supporting the use of population controls to increase power in these studies and the utility of using hospitalization as a proxy of severity. However, our results from the GRS analysis reported a need to maintain separated categories for severe-medium and critical illness.

We clearly replicated previously reported associations at 3p21.31 (near *SLC6A20* and *LZTFL1*-*FYCO1*) [7, 9, 22, 23] and 21q22.11 (in *IFNAR2*) [7, 9], and other findings in *ABO*, *OAS1*, *TYK2*, and *ARHGAP27* were validated. We also found a differential effect between males and females for SNPs in 3p21.31 and 21q22.11. Such differential genetic effects are also reflected in the heritability estimates. In this respect, the results strongly support that the genetic risk varies with sex and with a trend towards increasing differences with decreasing age, in agreement with the evidence suggesting a reduced impact of genetics with age [24]. While in the meta-analysis with other European studies the leading variants of 3p21.31 reached genome-wide significance in females, there was still a difference in effect size that, considering its magnitude, should not be disregarded. It is important to remark that these association signals found in males were not associated with the presence of comorbidities (see **Supplementary Figure 4**). In fact, genetic effects were only found for younger males (<60 years old), consistent with other studies [25] and strongly supporting those comorbidities outweigh genetic effects in disease outcomes in the older patients.

Some novel signals were found in our study, one in chromosome 19q13.12 (intergenic to *UPK1A* and *ZBTB32*, and also linked to the transcriptional regulation of *ARHGAP33*), and another in chromosome 9p13.3 (intergenic to *AQP7* and *AQP3*). Interestingly, we also found two sex-specific signals: *ELF5* in males and *TLE1* in females. *ELF5* has been recently reported as a new locus associated with critical illness in Europeans [26]. This locus reached genome-wide significance in our male meta-analysis of European cohorts (*p=*4.1×10^-8^). As regards of *TLE1*, even though this locus did not reach the standard genome-wide significance threshold (*p=*5.4×10^-7^), the signal is robust in the SCOURGE female GWAS. Given that the meta-analysis involved a low number of studies (and the top marker was not imputed in one of them), we believe this result should be taken with caution as further sex-specific studies will be needed to validate this finding.

*TLE1* encodes for the transducin-like enhancer of split 1, a co-repressor of other transcription factors and signalling pathways. Besides repressing the transcriptional activity of FOXA2 and of the Wnt signalling, TLE1 has been shown to negatively regulate NF-kB, which is fundamental in controlling inflammation and the immune response. The deficiency of TLE1 activity in mice results in enhancement of the NF-κB-mediated inflammatory response in diverse tissues including the lung [27]. Interestingly, TLE1 is one of the 332 high-confidence SARS-CoV-2 protein–human protein interactions identified so far [28]. Taken together, SARS-CoV-2 would be directly targeting the innate immunity and inflammation signalling pathways by interfering with the NF-κB activity. Thus, it is not surprising that TLE1 is a top-ranking regulator of inflammation that allows to transcriptionally distinguish mild from severe COVID-19 [29].

In the 19q13.12 locus, the most biologically plausible genes are *ARHGAP33* (also showing the best functional support based on the fine mapping variants) and *ZBTB32*. *ARHGAP33* is transcriptionally regulated by IRF1, a prominent antiviral effector and IFN-stimulated gene [30]. It also harbours NF-κB binding sites that modifies its expression in human lymphoblastoid cell lines by the presence of genetic variants in the binding site linked to many inflammatory and immune-related diseases including sepsis, and bacterial and viral infection [31]. Its expression is also altered in human induced pluripotent stem cells-derived pancreatic cultures in response to SARS-CoV-2 infection [32]. *ARHGAP33* was identified in an unbiased genome-wide CRISPR-based knockout screen in human Huh7.5.1 hepatoma cells infected by coronaviruses including SARS-CoV-2 and further interactome studies [33]. With respect to the transcription factor ZBTB32, it has been shown to impair antiviral immune responses by affecting cytokine production and the proliferation of natural killer and T cells, and the generation of memory cells [34]. In single cell studies, transcripts of *ZBTB32* were enriched in T follicular helper cells and were also expressed at significantly higher levels in hospitalized COVID-19 patients [35].

AQP3 is expressed most strongly in the kidney collecting duct, the gastrointestinal tract, large airways (in basal epithelial cells and the nasopharynx), skin, and the urinary bladder; while AQP7 is expressed primarily in the testis, fat cells and, to a lesser extent in a subsegment of the kidney proximal tubule [36]. In addition, AQP3 is upregulated in the lung tissues during viral or bacterial-induced diffuse alveolar damage [37]. Based on this, the evidence that SARS-CoV-2 viral proteins interacts with host proteins with the highest expression in lung tissues [38], and the functional evidence of the fine mapped variants in the GWAS being eQTLs in lung tissues, our data supports *AQP3* as the most likely gene of the 9p13.3 locus driving the association with COVID-19 hospitalization. Many patients develop acute respiratory distress syndrome (ARDS) during severe COVID-19 [39], and one of the hallmarks of ARDS is the increase of fluid volume (oedema) in the airspaces of the lung because of an increase in the alveolo-capillary membrane permeability. Some of the aquaporins, including AQP3, essentially function as water transport pores between the airways and the pulmonary capillaries [40], are key in lung fluid clearance and the formation of this lung oedema as a consequence of the lung injury [36]. In fact, the use of aquaporin modulators in lung inflammation and oedema has been proposed for potential use for the treatment of COVID-19 respiratory comorbidity [41].

We have also shown for the first time that COVID-19 severity risk suffers from ID, where individuals with higher levels of homozygosity associate with higher risk of being hospitalized and of developing severe COVID-19. Our results also suggested that autozygous rare recessive variants found in ROH across the genome, rather than homozygous common variants in strong LD, are underlying the ID. Furthermore, the ID is stronger in males than in females suffering from COVID-19 hospitalizations, especially in males ≥60 years old. Although these results may be found counterintuitive with the rest of findings, they are supported by the mutation accumulation senescence theory. Under this theory, alleles with detrimental effects that act in late life are expected to accumulate and cause senescence, thus increasing the ID [42]. We detected further sex-specific effects of homozygosity through ROHi. In hospitalized males, coagulation and complement pathways, which have been previously associated with severe COVID-19 [43], were enriched among the protein coding genes located in ROHi, highlighting the role of homozygosity whereas the Lectin pathway of complement activation is reported to be involved in the response to SARS-CoV-2 infection [44–46]. In hospitalized females, PI3K-Akt signalling genes were found to be enriched in ROH islands, whose networks are affected by a great variety of viruses [47].

Given that the effect of the genetic variants in SARS-CoV-2 severity is clearer among males and previous evidence on this regard, we elucubrate on the role of androgens in COVID-19 severity. Androgenic hormones have been suggested to be responsible of the excess male mortality observed in COVID-19 patients [48], and several lines of evidence suggest that the androgen receptor (AR) pathway is involved in the severity of SARS-CoV-2 infection: (1) A higher mortality rate among men has been established [49]; (2) A substantial proportion of individuals, both males and females, hospitalized for severe COVID-19 have androgenetic alopecia (AGA) [49]; (3) Most of the genes on COVID-19 severity in this study have been identified in male-only analyses, and these genes have been shown to interact with the AR. The following lines of evidence suggest the AR pathway is a mechanism responsible for some identified genes-COVID-19 severity relationship: (1) FYCO1 is regulated by the AR [50], and binding sites between the sex hormone receptor AR and FYCO1 have been demonstrated [50, 51]; (2) There is a cross-talk between the IFN pathways and the androgen signalling pathways [52], and androgens are regulated by IFNs in human prostate cells [53]; (3) *TMPRSS2*, another gene associated with COVID-19 severity in other studies, is induced by androgens through a distal AR binding enhancer [54]; (4) AR induces the expression of chemokine receptors such as CCR1; (5) Variants of *LZTFL1* gene are likely pathogenic for male reproductive system diseases [55]; (6) genetic polymorphisms in the AR (long polyQ alleles ≥23) and higher testosterone levels in subjects with AR long-polyQ appear to predispose some men to develop more severe disease [56]. Thus, it is not unexpected to find that antiandrogen treatments are under the focus as treatment options and prophylaxis of severe COVID-19 [49] and that randomized controlled clinical trials with bicalutamide (NCT04374279), degarelix (NCT04397718), and spironolactone (NCT04345887) are currently underway.

## Data Availability

Summary statistics of this study will be aggregated with those from the COVID-19 Host Genetics Initiative (https://www.covid19hg.org); data will also be shared upon request to the corresponding authors.

## Methods

### Recruitment of cases and phenotype definitions for the discovery phase

In Spain, 11,939 COVID-19 positive cases were recruited as part of SCOURGE study from 34 centres in 25 cities. The complete list of hospitals or research centers and the number of samples that each contributed to the study is shown in **Table S1**. Study samples and data were collected by the participating centers, through their respective biobanks after informed consent, with the approval of the respective Ethic and Scientific Committees. The whole project was approved by the Galician Ethical Committee Ref 2020/197. All samples and data were processed following normalized procedures. Study data were collected and managed using REDCap electronic data capture tools hosted at Centro de Investigación Biomédica en Red (CIBER) [57, 58] (**Supplementary Note**). Individuals were diagnosed as COVID-19 positive through a PCR-based test or according to local clinical and laboratory procedures. All cases were classified in a five-level severity scale (**Table 1**).

Two Spanish sample collections with unknown COVID-19 status were included as general population controls in some analyses: 3,437 samples from the Spanish DNA biobank (https://www.bancoadn.org) and 2,506 samples from the GR@CE consortium [17].

### Replication study

A total of 1,598 COVID-19 cases from six different Spanish Biobanks (Biobanco CNIO, Biobanco Vasco, Biobanco Hospital Ramón y Cajal, Biobanco Hospital Puerta de Hierro, Biobanco Hospital San Carlos, and Banco Nacional de ADN) were recruited under the ethical committee approval CEI PI 34_2020-v2. Additionally, 1,068 individuals from Spanish DNA biobank were included as controls in the analysis whenever necessary. We will refer to these cases and controls as the Centro Nacional de Investigaciones Oncológicas (CNIO) study.

### Genotyping

The discovery phase samples were genotyped with the Axiom Spain Biobank Array (Thermo Fisher Scientific) following the manufacturer’s instructions in the Santiago de Compostela Node of the National Genotyping Center (CeGen-ISCIII; http://www.usc.es/cegen). This array contains 757,836 markers, including rare variants selected in the Spanish population. Genomic DNA was obtained from peripheral blood and isolated using the Chemagic DNA Blood100 kit (PerkinElmer Chemagen Technologies GmbH), following the manufacturer’s recommendations.

For the replication study samples, a total of 250 ng of DNA was processed according to the Infinium HTS assay Protocol (Part # 15045738 Rev. A, Illumina), including amplification, fragmentation and hybridization using the Global Screening Array Multi-disease v3.0. This. This array contains a total of 730,059 markers and was scanned on an iScan platform (Illumina, Inc.). Clustering and genotype calling were performed using Genome Studio v2.0.4 (Illumina, Inc.).

### Quality control

A quality control (QC) procedure was carried out for the SCOURGE study samples and control datasets. First, a list of probe sets was removed based on poor cluster separation or excessive minor allele frequency (MAF) difference from The 1000 Genomes Project data (1KGP) [59]. Then, the following QC steps were applied using PLINK 1.9 [60] and a custom R script. We excluded variants with MAF<1%, call rate <98%, a difference in missing rate between cases and controls >0.02, or deviating from Hardy-Weinberg equilibrium (HWE) expectations (*p*<1×10^-6^ in controls, *p*<1×10^-10^ in cases, with a mid-*p* adjustment [61]). Samples with a call rate <98% and those in which heterozygosity rate deviated more than 5 SD from the mean heterozygosity of the study were also removed.

To assess kinship and assign ancestries, autosomal SNPs (MAF>5%) were pruned with PLINK using a window of 1,000 markers, a step size of 80 and a r^2^ of 0.1. Additionally, high-linkage disequilibrium (LD) regions described in Price et al. [62] were also excluded. A subset of 131,937 independent SNPs was used to evaluate kinship (IBD estimation) in PLINK. Given the possible confusion between relatedness and ancestry, we removed only one individual from each pair of individuals with PI_HAT>0.25 (second-degree relatives) that showed a Z0, Z1, and Z2 coherent pattern (according to theoretical expected values for each relatedness level). The unrelated SCOURGE individuals were merged with samples from 1KGP and the common SNPs were LD-pruned as previously indicated. Ancestry was then inferred with Admixture [63] using the defined 1KGP superpopulations. Those individuals with an estimated probability >80% of pertaining to European ancestry were defined as European (N=15,571).

Genomic principal components (PCs) were also computed using a LD-pruned (r^2^ < 0.1 with a window size of 1,000 markers) subset of genotyped SNPs passing quality check for controlling the population structure in the GWAS.

The CNIO study was filtered following the same QC procedures, where 220 individuals were identified as non-European and, therefore, were excluded from further analysis.

### Variant imputation

Imputation was conducted based on the TOPMed version r2 reference panel (GRCh38) [64] in the TOPMed Imputation Server. After post-imputation filtering (Rsq>0.3, HWE *p*>1×10^-6^, MAF>0.01), 15,045 individuals (9,371 COVID-19 positive cases and 5,674 population controls) and 8,933,154 genetic markers remained in the SCOURGE European study (discovery). The final dataset of the CNIO study (replication) included 2,446 individuals (1,378 COVID-19 positive cases and 1,068 population controls) and 8,895,721 markers. For directly genotyped variants, the original genotype was maintained in place of the imputed data.

### Statistical analysis

Association testing was computed by fitting logistic mixed regression models adjusting for age, sex, and the first 10 ancestry-specific PCs. SNPRelate [65] was used for prior LD-pruning and data management. Association analyses were performed in SAIGEgds [66], which implements the SAIGE [67] two-step mixed model methodology and the SPA test while using more efficient objects for genotype storage. A null model was fitted in the first step using the LD-pruned genotyped variants (MAF >0.005%, missing rate <98%) to estimate variance components and the GRM. Then, in a second step, association analyses were performed for both genotyped and imputed SNPs. Significance was established at *p*<5×10^-8^ after meta-analysis of the discovery and replication phases.

To align the results with those from the COVID-19 HGI, three outcomes were evaluated in relation to severity: hospitalization, severe COVID-19 (severity ≥3), and very severe COVID-19 (severity=4, critical illness). For each comparison, three control definitions were used (**Table 2**):

- A2 analysis: control samples from the general population.

- C analysis: COVID-19 positive not satisfying the case condition.

Additionally, the risk to COVID-19 infection was also analysed by comparing all COVID-19 positive cases with control samples from the general population.

All analyses were conducted for each complete dataset and stratified by sex and age (<60 years, ≥60 years). The SNP*sex (and SNP*age group) interaction term was tested for each SNP in the subset of clumped signals, adjusting the models for the same covariates.

Then, the main results of both Spanish cohorts (SCOURGE and CNIO) for the overall and sex-stratified analyses were meta-analysed assuming a fixed-effects model using METAL [68].

Because of the similarity of both the SCOURGE and CNIO studies in the clinical variables recorded and, more importantly, in the definition of the severity scale, the leading variants from the significant and validated loci in the hospitalization analysis were also analysed under a multinomial model (supplementary note).

### Validation of findings in independent studies

In order to validate the findings in independent study samples of European ancestry, a meta-analysis of hospitalization risk was performed for the overall and sex-stratified summary statistics of both Spanish cohorts (SCOURGE and CNIO) and other four sex-stratified Europeans studies from the COVID-19 HGI consortium (BelCOVID, GenCOVID, Hostage-Spain, and Hostage-Italy).

### Bayesian fine-mapping of GWAS findings

Credible sets were calculated for the GWAS loci to identify a subset of variants most likely containing the causal variant at 95% confidence level, assuming that there is a single causal variant and that it has been tested. We used *corrcoverage* for R [69] to calculate the posterior probabilities of the variant being causal for all variants with an r^2^>0.1 with the leading SNP and within 1 Mb. Variants were added to the credible set until the sum of the posterior probabilities was ≥0.95. VEP (https://www.ensembl.org/info/docs/tools/vep/index.html) and the V2G aggregate scoring from Open Targets Genetics (https://genetics.opentargets.org) were used to annotate the most prominent biological effects of the variants in the credible sets.

### Genetic risk score

A genetic risk score (GRS) was created for the SCOURGE cohort individuals and population controls using the list of SNPs associated with hospitalization, severity, or risk in the meta-analysis performed by the COVID-19 HGI [9]. We used the reported effects as weights and prioritized the hospitalization weight for variants significantly associated in the three analyses. To evaluate the existence of genetic risk differences along the disease stages, we fitted an ANOVA using the six-level severity scale (controls from the general population and the five severity levels defined in **Table 1**) as the independent variable. A *post hoc* Duncan test was performed to statistically assess the pairwise differentiation between the levels.

### SNP heritability of COVID-19 severity

We relied on GCTA-GREML 1.93.2beta [70] to assess the heritability of severe COVID-19 symptoms among SCOURGE patients, excluding those with cryptic relatedness or with missing genotypes above 0.5% and assuming a prevalence of COVID-19 hospitalization of 0.5%. This analysis considered all patients (modelling for age, sex, sex*age, and the 10 first PCs), and males and females separately (modelling for age and the 10 first PCs). We also partitioned the variance to assess the heritability changes among the patients <60 or ≥60 years old. We focused on the 547,206 autosomal variants with MAF>1% and missingness <0.5%. Assuming 0.5% of prevalence of severe COVID-19, and at least 1,500 cases and 1,500 controls per stratum, we estimate >97.9% power to detect a heritability >0.2.

### ROH calling

The ROH segments longer than 300 Kb were called in SCOURGE using PLINK 1.9 in the European QC-ed genotyped dataset (before imputation) with the following parameters: *homozyg-snp 30*, *homozyg-kb 300*, *homozyg-density 30*, *homozyg-window-sn 30, homozyg-gap 1000, homozyg-window-het 1, homozyg-window-missing 5*, *homozyg-window-threshold 0.05.* No LD pruning was performed.

### Calculating genomic inbreeding coefficients

Different genomic inbreeding coefficients were calculated [71]:

**F_ROH_** measures the actual proportion of the autosomal genome that is autozygous above a specific minimum length ROH threshold.

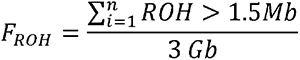

**F_GRM_** is an alternative genomic inbreeding coefficient that was obtained using PLINK’s parameter -ibc (Fhat3). This coefficient described by Yang et al. 2011 [70]; where *N* is the number of SNPs, *p*_i_ is the reference allele frequency of the *i*th SNP, and *x*_i_ is the number of copies of the reference allele. The reference allele frequencies were site-specific and included only variants with MAF >0.05.

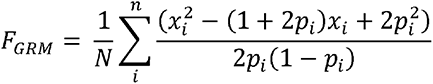

### Testing and replicating the inbreeding depression

Inbreeding depression (ID) is defined as the change in the mean phenotypic value in a population because of inbreeding [12, 13]. The ID was modelled in SCOURGE by a multiple logistic regression. The covariables used in this study were sex, age, and the first ten PCs.

The results were replicated in a cohort gathered by Tomoko et al. 2021 [24]. This consists of clinical and genomic data from 4,418 European ancestry individuals (3,946 hospitalized COVID-19 cases and 422 controls): 2,597 men (1,072 men <60 years old, 1,525 men ≥60 years old) and 1,821 women (808 <60 years old, 1,013 women ≥60 years old). The cohort was built by harmonizing individual-level data from 16 different studies [24]. The F_ROH_ and F_GRM_ coefficients were obtained following the procedure explained above. The model described above with the same covariables (age, sex, and the first then PCs) was applied in this cohort.

Genome-specific effects on COVID-19 severity and hospitalization were tested through ID in genomic windows, ROH islands (ROHi) and regions of heterozygosity (RHZ) (**Supplementary Note**).

## Acknowledgements

This study has been funded by Instituto de Salud Carlos III (COV20_00622 to A.C., COV20/00792 to M.B, COV20_00181 to J.M.A.G., COV20_1144 to M.A.J.S., PI20/00876 to C.F) and cofunded by European Union (ERDF) “A way of making Europe”. Fundación Amancio Ortega, Banco de Santander (to A.C), Estrella de Levante S.A. and Colabora Mujer Association (to EG-N) and Obra Social La Caixa (to R.B); Agencia Estatal de Investigación (RTC-2017-6471-1 to C.F), Cabildo Insular de Tenerife (CGIEU0000219140 “Apuestas científicas del ITER para colaborar en la lucha contra la COVID-19” to C.F., have also contributed to its funding.

R.L-R is granted by Cátedra de Medicina Genómica IIS-Fundación Jiménez Díaz-UAM, M.B. by Nextgeneration EU funds. M.C., P.M., M.A.J.S., A.F.R. are granted by the Miguel Servet Programme (CP17/00006, CP16/00116, CPII20CIII/0001, CPII20CIII/0001 respectively) and B.A. by the Juan Rodés Programme (JR17/00020), all of them from Instituto de Salud Carlos III, and cofunded by European Union (ERDF) “A way of making Europe”.

The contribution of the Centro National de Genotipado (CEGEN), and Centro de Supercomputación de Galicia (CESGA) for funding this project by providing supercomputing infrastructures, is also ackowledged. Authors are also particularly grateful for the supply of material and the collaboration of patients, health professionals from participating centers and biobanks. Namely Biobanc-Mur, and biobancs of the Complexo Hospitalario Universitario de A Coruña, Complexo Hospitalario Universitario de Santiago, Hospital Clínico San Carlos, Hospital La Fe, Hospital Universitario Puerta de Hierro Majadahonda - Instituto de Investigación Sanitaria Puerta de Hierro - Segovia de Arana, Hospital Ramón y Cajal, IDIBGI, IdISBa, IIS Biocruces Bizkaia, IIS Galicia Sur. Also biobanks of the Sistema de Salud de Aragón, Sistema Sanitario Público de Andalucía, and Banco Nacional de ADN.

---

**Extended Data Table 1.**
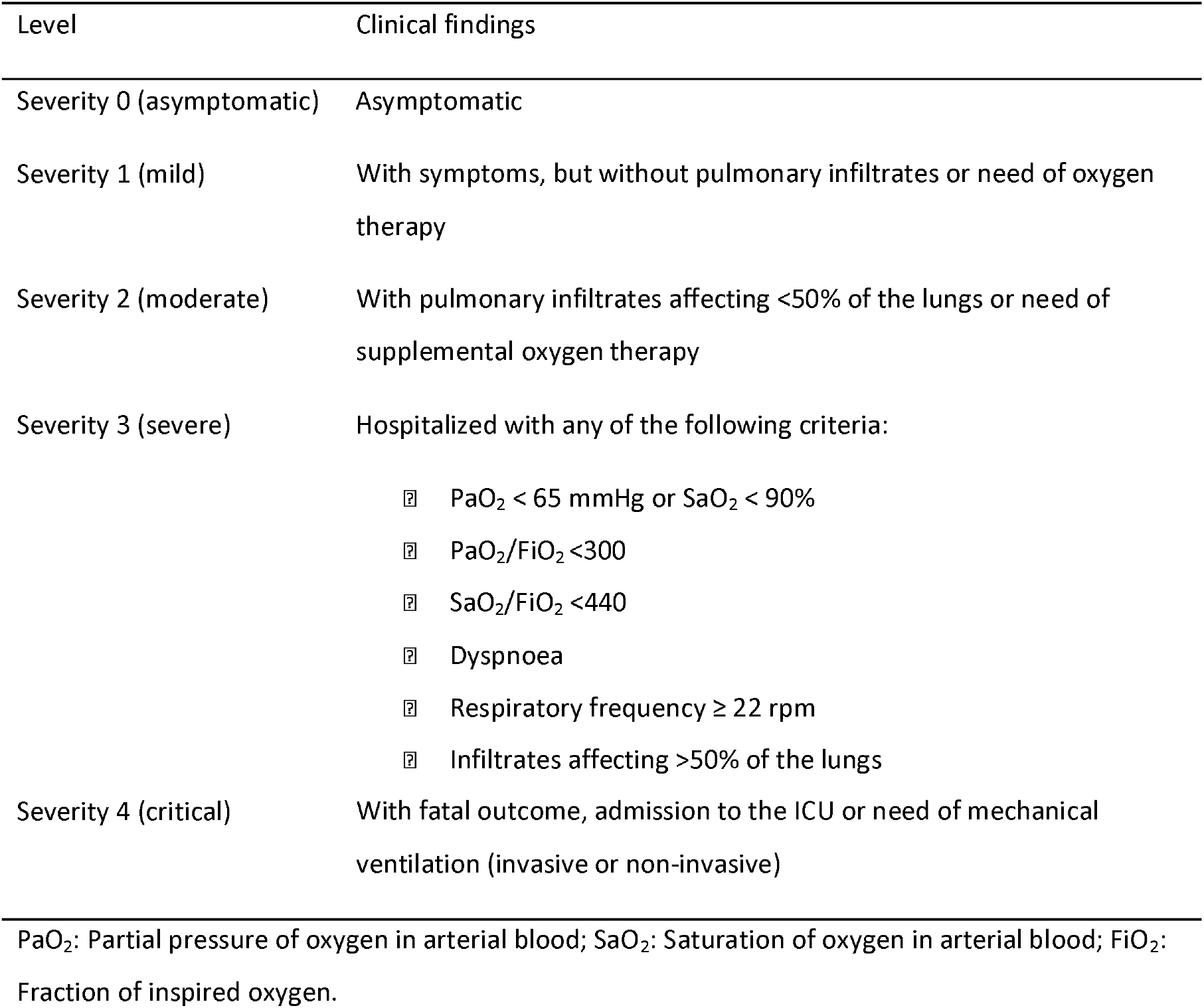
Five-level severity scale used to classify SCOURGE patients.

| Level | Clinical findings |
| --- | --- |
| Severity 0 (asymptomatic) | Asymptomatic |
| Severity 1 (mild) | With symptoms, but without pulmonary infiltrates or need of oxygen therapy |
| Severity 2 (moderate) | With pulmonary infiltrates affecting <50% of the lungs or need of supplemental oxygen therapy |
| Severity 3 (severe) | Hospitalized with any of the following criteria:<br>□ $\text{PaO}_2 < 65 \text{ mmHg}$ or $\text{SaO}_2 < 90\%$<br>□ $\text{PaO}_2/\text{FiO}_2 < 300$<br>□ $\text{SaO}_2/\text{FiO}_2 < 440$<br>□ Dyspnoea<br>□ Respiratory frequency $\geq 22 \text{ rpm}$<br>□ Infiltrates affecting >50% of the lungs |
| Severity 4 (critical) | With fatal outcome, admission to the ICU or need of mechanical ventilation (invasive or non-invasive) |

$\text{PaO}_2$ : Partial pressure of oxygen in arterial blood; $\text{SaO}_2$ : Saturation of oxygen in arterial blood; $\text{FiO}_2$ : Fraction of inspired oxygen.

**Extended Data Table 2.** Baseline characteristics of European patients from SCOURGE included in the analysis.

| Variable | Global<br>N = 9,371 | males<br>N = 4,343 | females<br>N = 5,028 |
| --- | --- | --- | --- |
| Age – mean years (SD) | 62.6 (17.9) | 64.3 (16.3) | 61.1 (19.1) |
| Severity - N (%) |  |  |  |
| 0 - asymptomatic | 582 (6.6) | 161 (3.9) | 421 (8.9) |
| 1 - mild | 2,689 (30.3) | 748 (18.2) | 1,941 (40.8) |
| 2 - intermediate | 2,099 (23.6) | 1,093 (26.5) | 1,006 (21.1) |
| 3 - severe | 2,379 (26.8) | 1,300 (31.6) | 1,079 (22.7) |
| 4 - critical illness | 1,128 (12.7) | 817 (19.8) | 311 (6.5) |
| Hospitalization - N (%) | 5,966 (63.8) | 3,436 (79.3) | 2,530 (50.5) |
| Severe COVID-19 - N (%) | 3,507 (39.2) | 2,117 (51.2) | 1,390 (28.9) |
| Critical illness - N (%) | 1,128 (12.6) | 817 (19.8) | 311 (6.5) |
| Comorbidities - N (%) |  |  |  |
| Vascular/endocrinological | 4,099 (43.7) | 2,207 (50.8) | 1,892 (37.6) |
| Cardiac | 1,057 (11.3) | 634 (14.6) | 423 (8.4) |
| Nervous | 773 (8.3) | 341 (7.9) | 432 (8.6) |
| Digestive | 264 (2.8) | 153 (3.5) | 111 (2.2) |
| Onco-hematological | 647 (6.9) | 411 (9.5) | 236 (4.7) |
| Respiratory | 905 (9.7) | 565 (13.0) | 340 (6.8) |

**Extended Data Figure 1.**
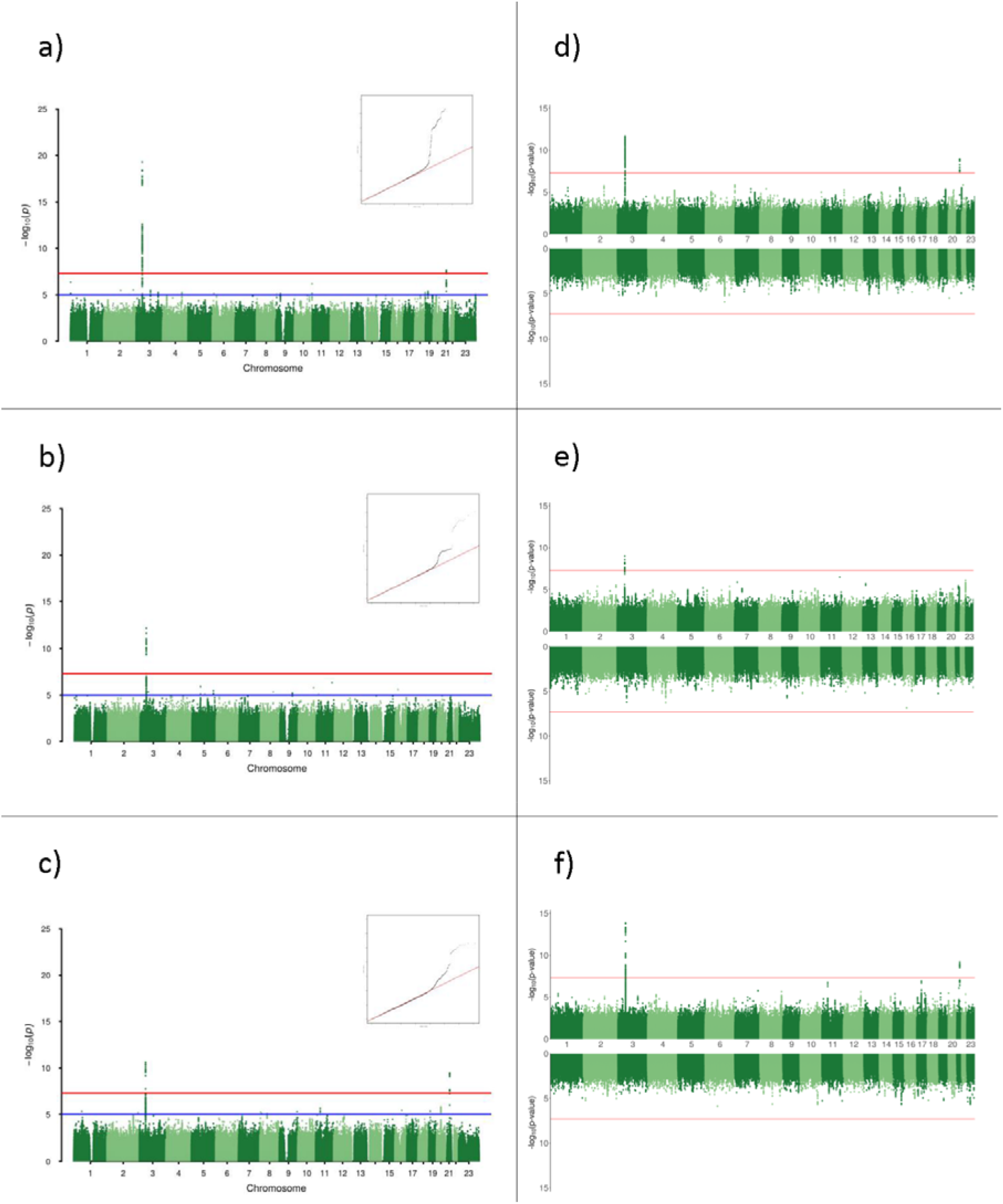
Manhattan plots and quantile-quantile plots of the GWAS results of the A1 analysis from the overall SCOURGE study and Miami plots for the sex-stratified analysis (top: males, bottom: females). a, b, c: Manhattan plots for severe illness, critical illness, and risk of infection, respectively. d, e, f: Miami plots for sex-stratified analyses in severe illness, critical illness, and risk of infection, respectively.

**Extended Data Figure 2.**
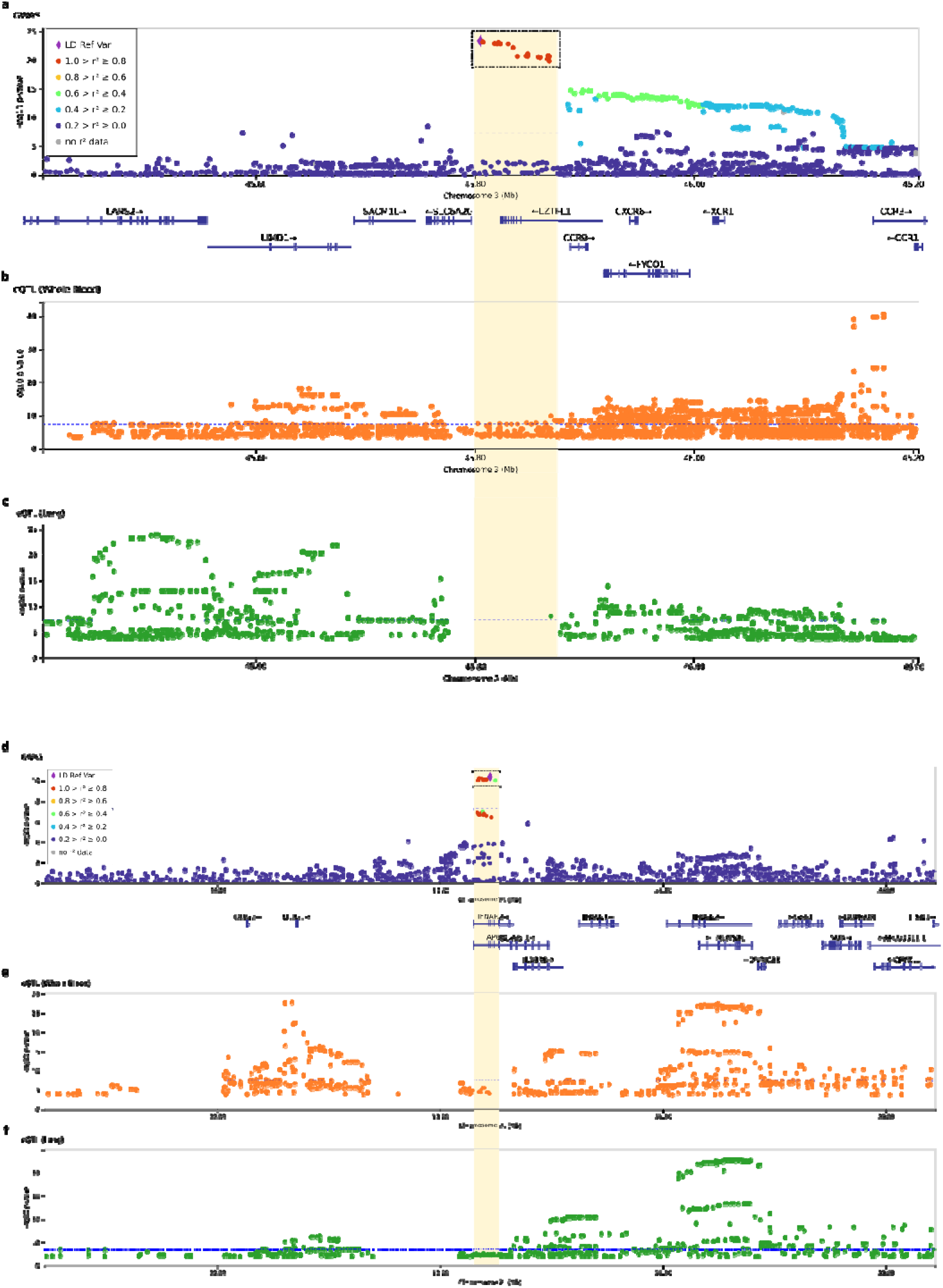
Regional plots of two previously reported association signals in 3p21.31 (a-c) and 21q22.11 (d-f). The x axes reflect the chromosomal position, and the y axes the -log(p-value) in the SCOURGE study. On panels a) and d), the sentinel variant is indicated by a diamond and all other variants are colour coded by their degree of linkage disequilibrium with the sentinel variant in Europeans. Credible sets for each signal are shown by squares. The horizontal dotted blue line corresponds to the threshold for genome-wide significance (*p*=5×10^-8^). In the rest of panels, the x axes reflect the chromosomal position, and the y axes the -log(p-value) resulting from the eQTL analyses in whole blood (b and e) and in the lung (c and f) whenever a significant finding is available from GTEx v8.

## Supplementary Material

**Supplementary Figure 1.**
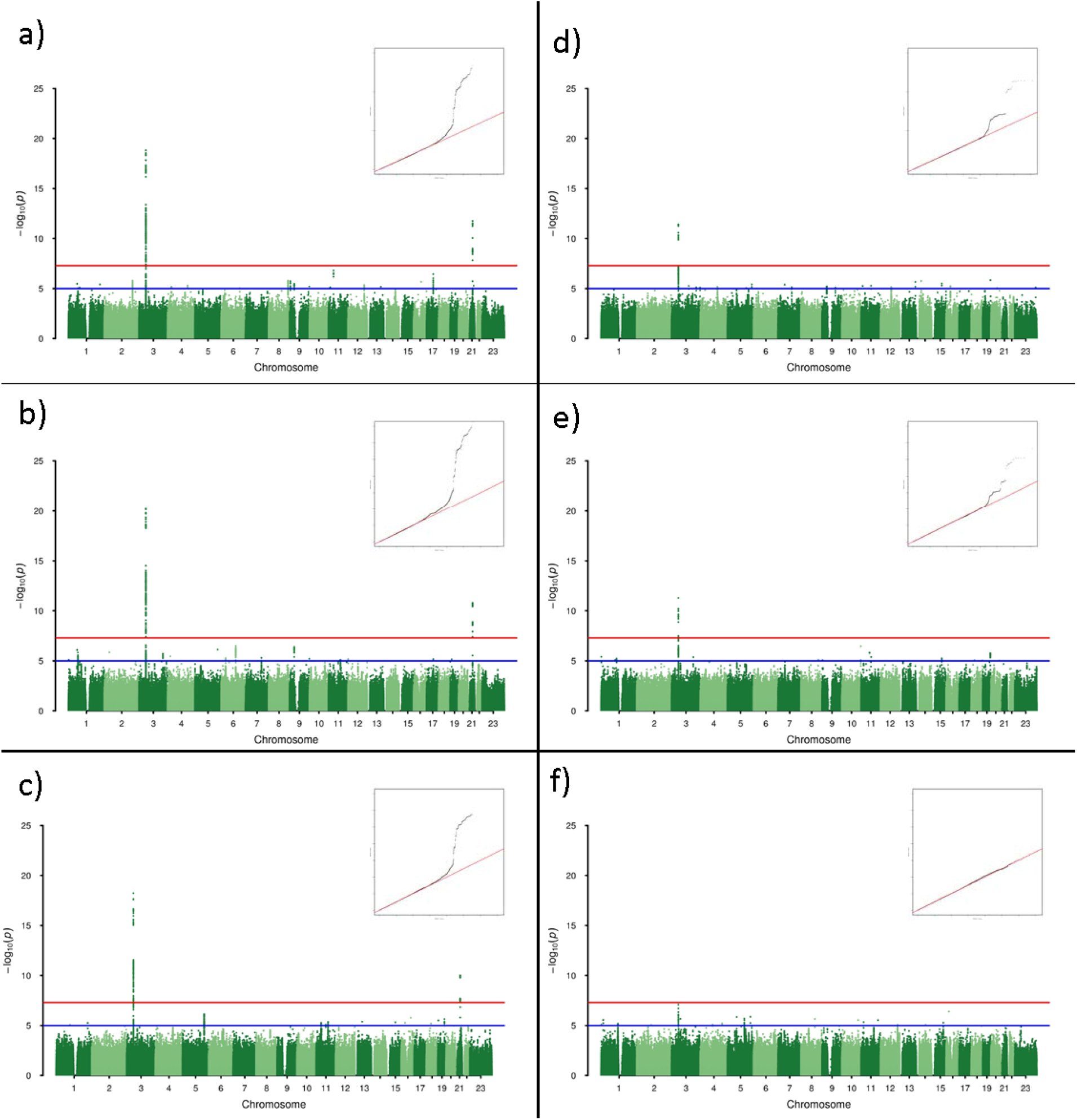
Manhattan plots and quantile-quantile plots of the GWAS results for the overall SCOURGE study corresponding to A2 (left) and C (right) analyses for hospitalization (a, d), severe illness (b, e), and critical illness (c, f).

**Supplementary Figure 2.**
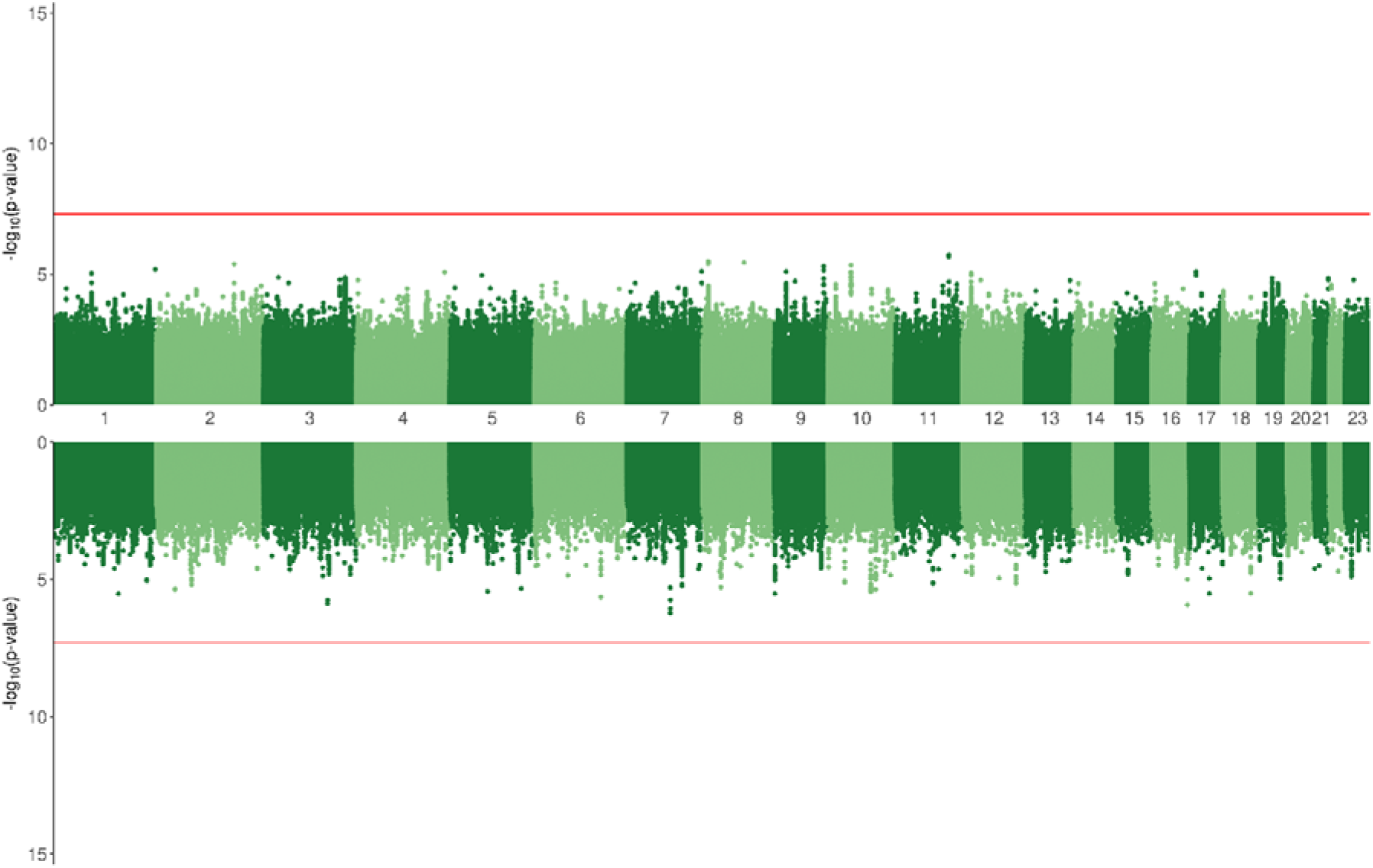
Miami plot of the GWAS results of SCOURGE for sex-disaggregated analyses of the presence of comorbidities. Top: males; bottom: females

**Supplementary Figure 3.**
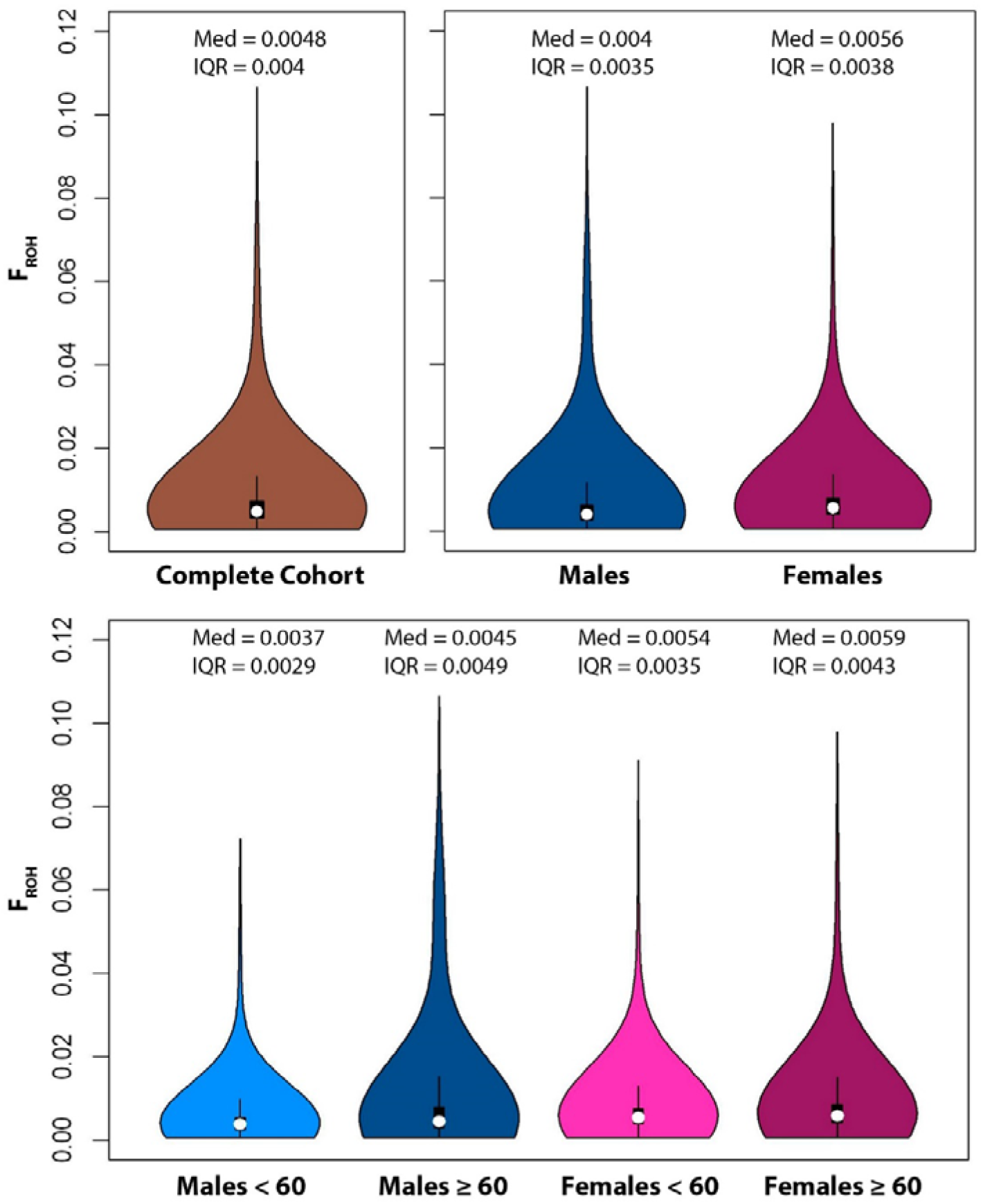
Violin plots showing the distribution of ROH longer than 1.5 Mb for different population groups in the SCOURGE study. Median and interquartile range are shown for each group.

**Supplementary Figure 4.**
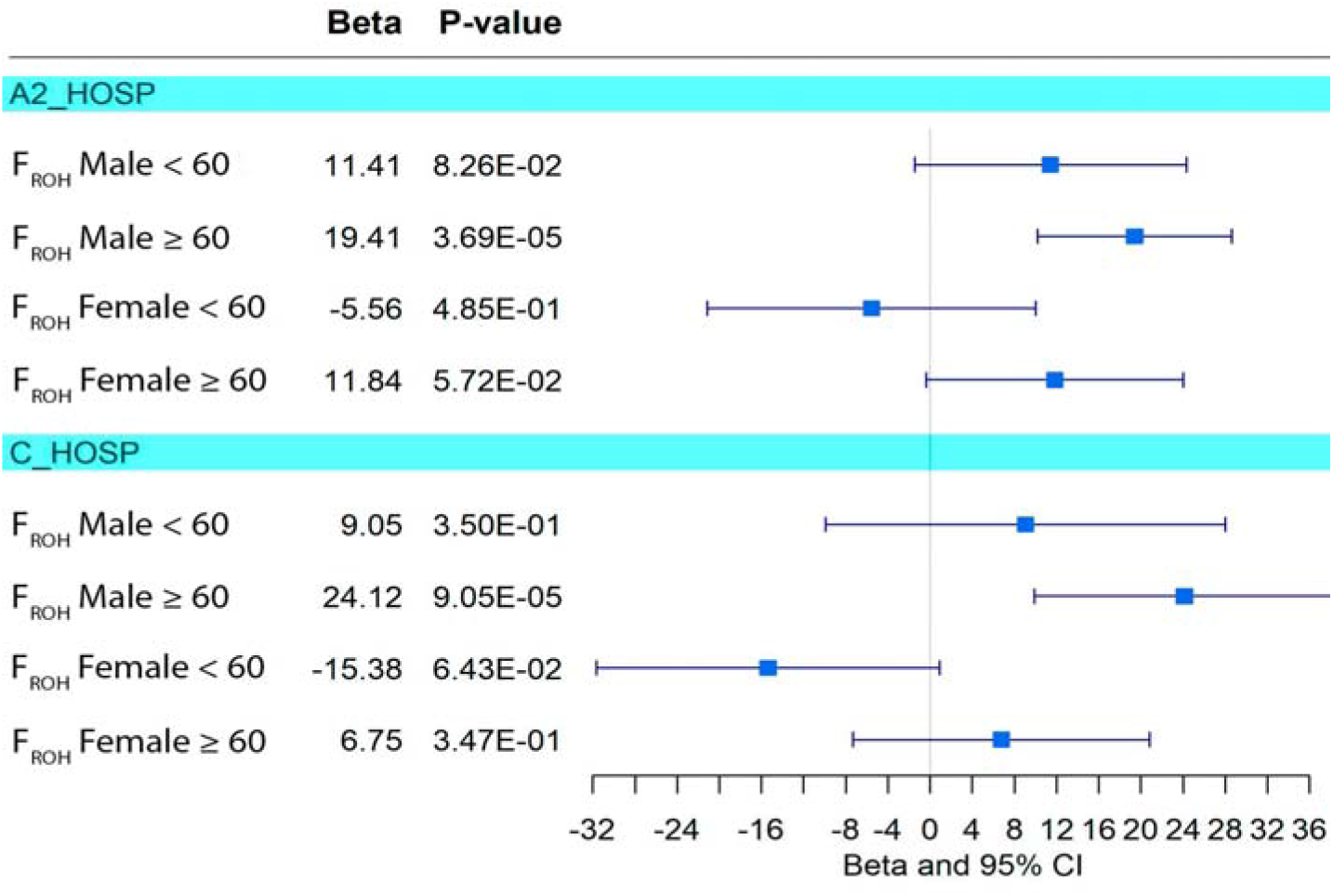
Effect of the inbreeding depression on COVID 19 hospitalization using different control groups. Two different population groups were used as control group: 1) Healthy COVID-19 negative individuals, and 2) Non-hospitalized COVID-19 positive individuals. Forest plots are shown for individuals disaggregated by sex and age.

**Supplementary Figure 5.**
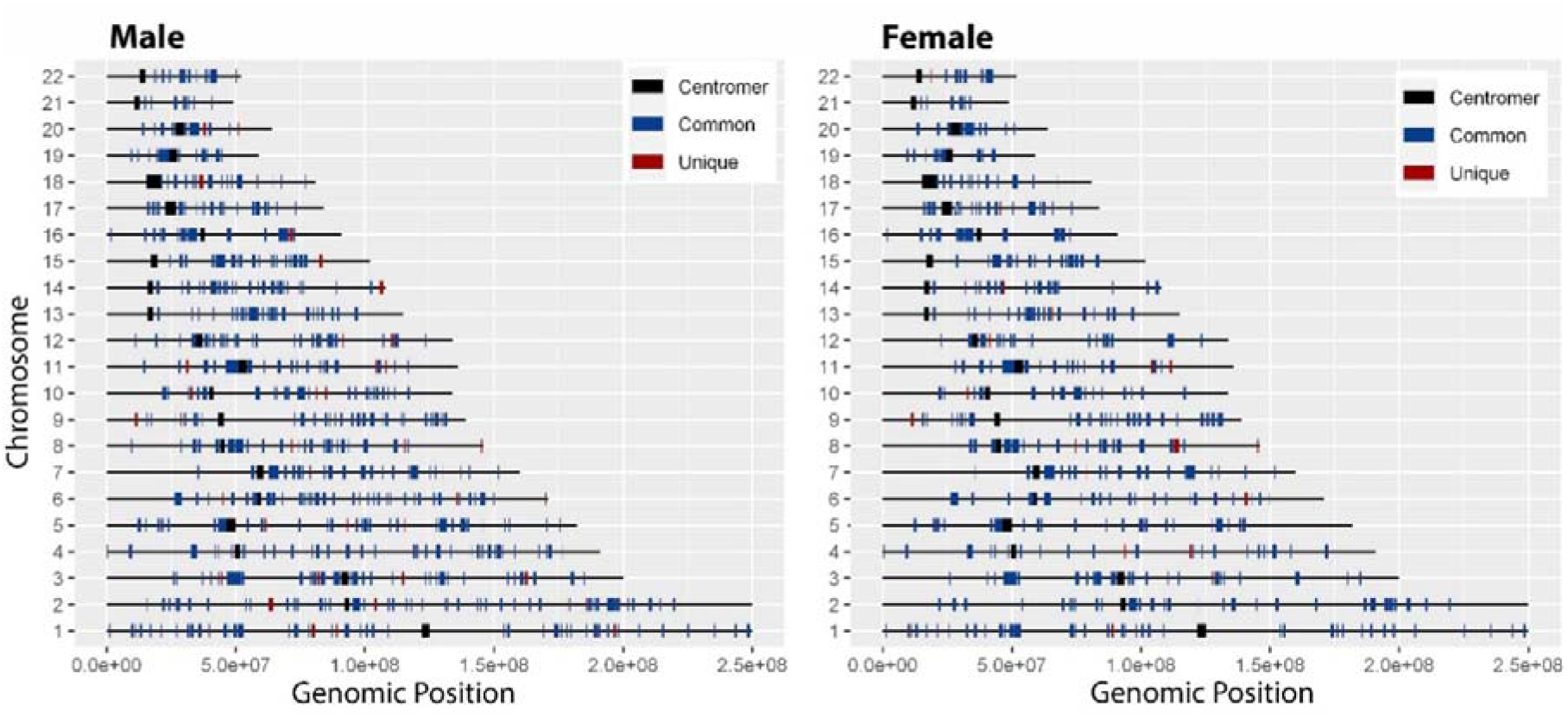
Genomic representation of the chromosomal location and size of the runs of homozygosity islands (ROHi) for hospitalized males and females in the SCOURGE study. Unique ROHi of hospitalized males and females are shown in red. Common ROHi between hospitalized and non-hospitalized individuals are shown in blue.

**Supplementary Figure 6.**
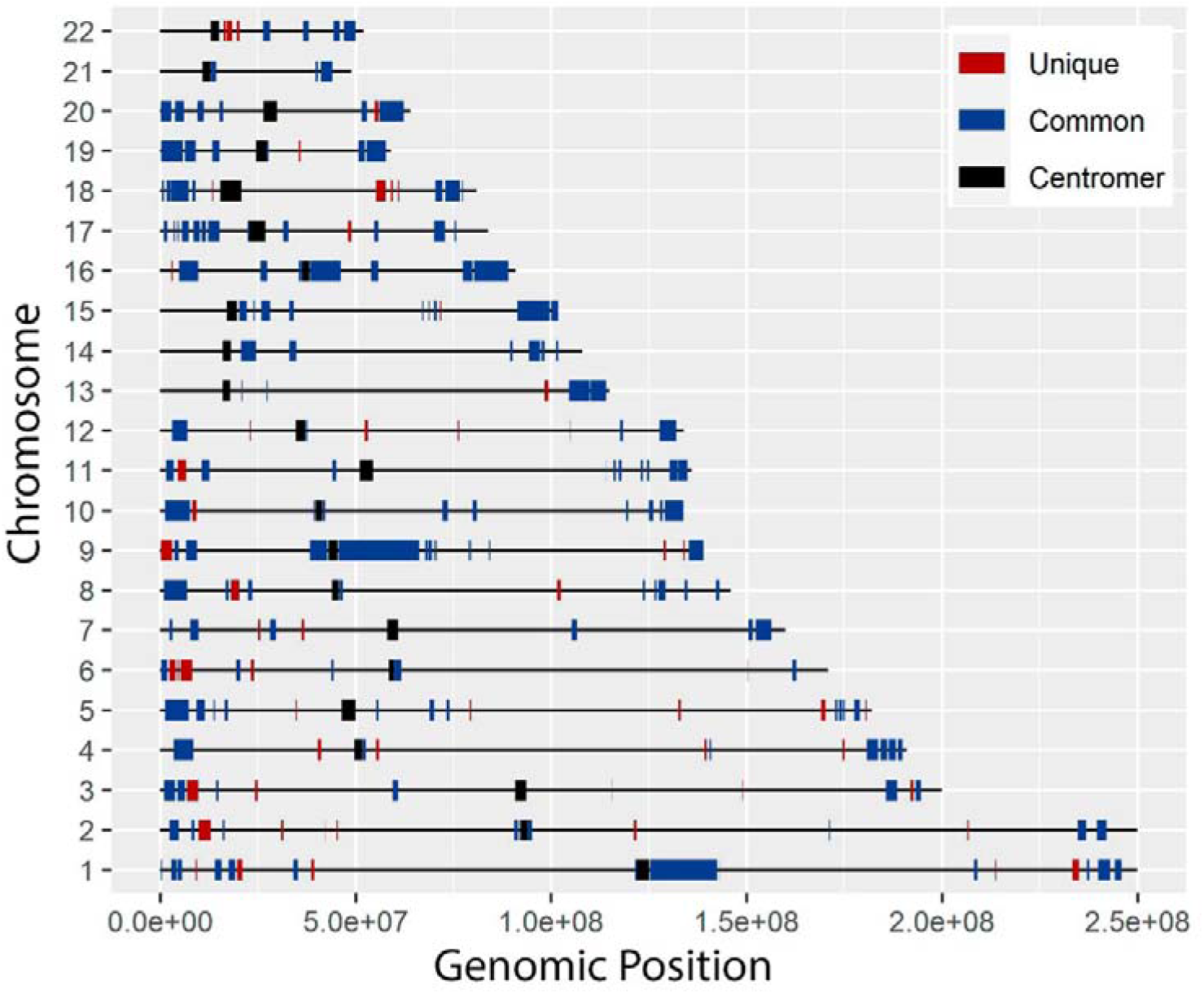
Genomic distribution of regions of heterozygosity (RHZ). Genomic representation of the chromosomal location and size of regions of heterozygosity for non-hospitalized males. Unique RHZ of non-hospitalized males are shown in red. Common ROHi between hospitalized and non-hospitalized males are shown in blue.

**Supplementary Tables** are provided by a separate excel file

## Supplementary Note

### Research electronic data capture (REDCap)

REDCap tools, hosted at Centro de Investigación Biomédica en Red (CIBER), was used to collect and manage the demographic, epidemiological, and clinical variables, together with the results of laboratory tests and imaging studies.

REDCap is a secure, web-based software platform designed to support data capture for research studies, providing 1) an intuitive interface for validated data capture; 2) audit trails for tracking data manipulation and export procedures; 3) automated export procedures for seamless data downloads to common statistical packages; and 4) procedures for data integration and interoperability with external sources.

### Multinomial regression on severity scale

As the GRS analysis outlined the existence of three severity categories in the SCOURGE study, as opposed to the clinically-based six-level scale, we used the multinomial model to test the association of this three-level severity scale (“mild”: control+asymptomatic+mild severity level; “intermediate”: intermediate+severe cases; “severe”: very severe cases) with the clumped loci that reached genome-wide significance in the meta-analysis of SCOURGE and CNIO studies (Table 1). Multinomial regressions were performed with the *mlogit* R library [1]. The null hypothesis for the leading variants was tested with the likelihood-ratio test. **Supplementary** Table 6 shows the results of multinomial regression for both the SCOURGE and CNIO studies. The SNPs showing a p-value < 0.0056 (Bonferroni adjusted threshold of 0.05/9) were considered significant. All variants remained significantly associated with the phenotype in the SCOURGE study, yet only four variants (three in 3p21.31 and the one in 9p13.3) were significantly associated in the CNIO cohort.

### Evaluating the associations of leading SNPs in relation with comorbidities

Further analyses were carried on hospitalized patients from the SCOURGE study to exclude a confounder effect of comorbidities in the genetic associations reported in this study. Firstly, we performed sex-disaggregated GWAS analyses on the presence/absence of comorbidities. No genome-wide significant associations were found, concluding that there is no evidence of direct association of comorbidities with the reported sex-specific signals (see **Supplementary Figure 2**).

Additionally, we adjusted the logistic models by the comorbidities of **Extended Data Table 2** (vascular, cardiac, nervous, digestive, onco-haematological, or respiratory) for each of the leading variants depicted in Table 1 and Table 2, adjusting also for age, sex, and 10 PCs. This confirmed that none of the leading variants was individually associated with any of the comorbidities recorded. Besides this, we also confirmed that there was a lack of confounding with the most frequent specific comorbidities (arterial hypertension, hypercholesterolemia, diabetes, EPOC or other chronic respiratory diseases, and obesity).

### Measuring genome-specific effects on COVID-19 severity and hospitalization

Different approaches were used to learn more about the genetic architecture of COVID-19 severity, namely the assessment of inbreeding depression (ID) in genomic windows, of the islands of runs of homozygosity (ROHi), and of the regions of heterozygosity (RHZ).

First, region-dependent ID was tested in the SCOURGE study by assessing the association of hospitalization and severity with ROH in nearly a thousand 3 Mb-wide windows along the genome (significance established at *p*<5×10^-5^ after Bonferroni correction). We found no evidence of major loci that may be exerting large effects, rather the ID was polygenetic in origin.

ROHi are defined as regions in the genome where the proportion of individuals of a population deviates from the expected under a binomial distribution. These regions have been found to be enriched with protein coding genes under selection [2, 3]. To search for ROHi in the SCOURGE study, a sliding window of 100 kb was used. In every 100 kb genomic window, the number of subjects with ROH was obtained and a binomial test was applied (threshold for significance established at *p*<2×10^-5^, corresponding to an adjustment for 2,500 windows). To prevent sampling bias, a resampling approach was followed. ROH from 100 men and women separately in both hospitalized and non-hospitalized groups were resampled (with replacement) 500 times and each replicate followed the above indicated methodology. Lastly, consecutive windows found to be statistically significant in at least 400 resampling events were considered as a part of the same ROHi. It was considered that both groups had the same specific ROHi if they shared ³ 75% of their genomic positions. Protein coding genes present in the ROHi were obtained using the *biomaRt* R package and Ensembl database and an enrichment pathway analysis was done on the gene lists using g:Profiler (https://biit.cs.ut.ee/gprofiler/gost, last access: August 23 2021). We found 592 ROHi in hospitalized males, 38 of them (6.4%) were unique to this group and were not found in non-hospitalized males (**Supplementary Figure 5**, **Supplementary Table 11**). A total of 152 protein coding genes were present in those 38 unique ROHi. In **Supplementary Table 12** we show an enrichment analysis of pathways based on those 152 protein-coding genes, strikingly revealing links with coagulation and complement pathways. Different ROHi were found to be unique for hospitalized COVID-19 females (**Supplementary Figure 5**, **Supplementary Table 11**). From a total of 433, 19 unique ROHi with 44 protein-coding genes were found in hospitalized females. Instead of coagulation or the complement, other pathways were enriched among females (**Supplementary Table 12**).

Finally, we searched for RHZ, where ROH are scarce or absent. To search for RHZ, an extra step of QC consisting of removing the SNPs in LD using PLINK was performed before calling for ROH. ROH longer than 100 Kb were called for this analysis and a 100 Kb sliding window was used. Two different cut-offs were considered to call RHZ in each window: a) No individual is homozygous, or b) 2% or fewer of the individuals are homozygous. Consecutive windows that fulfilled this requirement were considered part of the same RHZ. Among males, we found a total of 239 RHZ in non-hospitalized control group (non-hospitalized COVID-19 and population controls) and 214 RHZ in hospitalized COVID-19 patients. A total of 61 of the RHZ present in non-hospitalized individuals were found to be unique of this group (**Supplementary Figure 6**, **Supplementary Table 11**). Unique RHZ in non-hospitalized COVID-19 patients involved a total of 707 protein-coding genes. A total of 33 pathways were significantly enriched from this gene list, being olfactory receptor activity and sensory perception of smell the most significant ones (**Supplementary Table 12**). Surprisingly, we found 36 RHZ, where no individual has ROH, in both hospitalized and non-hospitalized COVID-19 individuals. These 36 RHZ involved 67 genes related to olfactory receptors, spermatogenesis, and survival of motor neurons.

